# Greater sustained sensorimotor function recovery and neuropathic pain reduction with Cognitive Multisensory Rehabilitation compared to adaptive fitness in adults with spinal cord injury: a pilot clinical trial

**DOI:** 10.64898/2026.03.26.26349257

**Authors:** Ann Van de Winckel, Sydney T. Carpentier, Sara Bottale, Jena Blackwood, Wei Deng, Lin Zhang, Timothy J. Hendrickson, Bryon A. Mueller, Ruhollah Nourian, Sheri Melander-Smith, Leslie R. Morse, Kelvin O. Lim

## Abstract

**Introduction:** Adults with spinal cord injury (SCI) often experience reduced or lost sensation and movement, impairing the ability of the brain to locate paralyzed body parts, which, in turn, compromises sensorimotor recovery. This disruption of the internal body map of the brain, or mental body representations (MBR), also contributes to neuropathic pain in about 69% of adults with SCI. Medications for neuropathic pain are often ineffective and can cause adverse reactions. Our previous pilot clinical trial showed that Cognitive Multisensory Rehabilitation (CMR), a physical therapy that restores MBR, produced significant, lasting reductions in neuropathic pain, improved sensorimotor function, and enhanced brain function. Building on these results, we examined whether 8 weeks of CMR or adaptive fitness (1) improved sensorimotor function and reduced pain; (2) greater brain activity and connectivity related to sensorimotor function and MBR in adults with SCI.

**Methods:** Sixteen participants (52+/-8 years old, 13+/-10 years post-SCI) were randomized to 8 weeks of CMR or adaptive fitness (45 min, 3x/week). Ten participants had neuropathic pain of 3/10 or greater. Pain and sensorimotor function were assessed at baseline, post-intervention, and 3-month follow-up using the Numeric Pain Rating Scale (NPRS), ASIA Impairment Scale (AIS), and Neuromuscular Recovery Scale (NRS). Functional MRI included resting-state and 4 tasks: imagining feeling the left leg, imagining moving the left leg, whole-body movement imagery, and a sensation task.

**Results:** After CMR, participants improved on AIS with large effect sizes (touch: d=1.54; pinprick: d=1.83; lower limb motor function: d=1.32), while adaptive fitness had small/moderate effects (touch: d=0.49; pinprick: d=0.53; lower limb motor function: d=0.74). CMR also showed larger effect sizes for NRS (core: d=2.19; upper limb: d=0.69; lower limb: d=0.74) than fitness (core: d=0.73; upper limb: d=0.34; lower limb: d=0.00). Benefits persisted at follow-up. Highest neuropathic pain intensity reduced post-CMR and at 3-month follow-up (d=0.48; d=0.63). Pain increased slightly after fitness (n=6; d=-0.19; d=-0.41). CMR increased brain connectivity and activation during the leg imagery task. Increased activation during whole-body imagery was greater after CMR than fitness.

**Discussion:** These preliminary results support the potential of CMR to improve function and reduce neuropathic pain in adults with SCI, warranting larger confirmatory trials.

**Clinicaltrial.gov**: NCT05167032

## 1 Introduction

Spinal cord injury (SCI) imposes a severe dual burden on individuals, characterized not only by profound sensorimotor deficits but also by debilitating neuropathic pain, affecting about 69% of this population.(1–4) Traditional rehabilitation focuses heavily on functional motor recovery. Concurrent management of neuropathic pain is often reliant on pharmacological treatments that are frequently insufficient and can often cause significant adverse reactions. Consequently, there is an urgent need for effective, non-pharmacological interventions that address both functional recovery and pain to improve quality of life for adults with SCI.

A critical, yet often overlooked, driver of both poor sensorimotor recovery and neuropathic pain is the disruption of the brain’s ‘mental body representations’ (MBR). MBR encompasses whole-body awareness, and the brain’s ability to locate body parts in space via visuospatial body maps.(3–7) When SCI interrupts sensory and motor signals, the brain areas responsible for these maps—specifically the parietal operculum and insula for body awareness, the posterior parietal cortex for visuospatial body maps within the sensorimotor network, and the posterior cingulate cortex for self-location and body ownership—lose their normal input.(8–11)

We have previously demonstrated that SCI leads to weaker sensorimotor network connectivity and maladaptive neuroplasticity, compared to non-injured adults.(9) Without accurate MBR, the brain cannot locate the (paralyzed) limbs, thus, severely limiting motor recovery (e.g., walking), even when residual muscle strength exists, because the brain does not know how or where to activate those muscles. This same maladaptive cortical reorganization is a well-documented contributor to neuropathic pain onset and maintenance.(3,4,12,13) As the parietal operculum and insula are key regions for both MBR and pain,(14,15) restoring MBR is expected to improve sensorimotor recovery and reduce neuropathic pain.

Our previous pilot clinical trial demonstrated that Cognitive Multisensory Rehabilitation (CMR)─a physical therapy approach designed specifically to restore MBR─ holds significant promise, increasing brain activity and connectivity in areas relevant for MBR and sensorimotor function.(10,16) The clinical sensorimotor recovery after CMR is consistent with other studies in stroke populations.(8,17–20) Our earlier case studies further showed that CMR helped restore vision, sensory, and motor function in a person with cortical blindness and quadriplegia, and, relieved phantom limb pain after amputation.(21,22)

Based on these findings, we investigated whether 8 weeks of CMR could yield greater sensorimotor recovery than an active control intervention. We selected adaptive fitness as the control because it is a standard, community-accessible exercise program, commonly provided to adults with SCI following clinical neurorehabilitation. While systematic reviews show that fitness training improves cardiovascular health and upper limb strength, these programs, excluding interventions needing more hands-on assistance like body-weight supported treadmill training, typically focus on non-paretic limbs and do not explicitly target MBR restoration or neuropathic pain relief.(23–25).

Because MBR disruption occurs regardless of pain status, we recruited a broad SCI cohort to target this underlying mechanism for sensorimotor recovery, rather than restricting inclusion to those with neuropathic pain. Still, we prospectively tracked neuropathic pain as an important secondary outcome. We hypothesized that CMR would restore MBR and thereby reduce neuropathic pain in participants presenting with it. We therefore aimed to determine if 8 weeks of CMR or adaptive fitness (1) improved sensory and motor function (primary) and reduced neuropathic pain when present (secondary), and (2) enhanced brain activity and connectivity related to sensorimotor function and body awareness.

## 2 Materials and methods

### 2.1 Study design

Participants were recruited for this pilot randomized controlled trial between 6/2/2022 and 7/27/2023. The study followed the Declaration of Helsinki’s principles (2013).(26) The University of Minnesota (UMN)’s Institutional Review Board approved the study (STUDY00014710) on 2/28/2022.

### 2.2 Recruitment

Participants were recruited via fliers in the Paralyzed Veterans of America newsletter, the Minneapolis Veterans Affairs (VA) Medical Center, clinics and rehabilitation centers in the Twin Cities as part of the Minnesota Regional Spinal Cord Injury Model System, and clinicaltrial.gov (NCT05167032).

### 2.3 Participants

Participants were included if they had an incomplete or complete SCI at least 3 months ago and were medically stable. Participants were excluded if they had a ventilator dependency, uncontrolled seizure disorder, severe pressure sores hindering them from sitting for 45 min, cognitive and/or communicative disability (e.g., due to brain injury) precluding learning or following directions, other major medical complications, or pregnancy (current or planned). Participants with MRI contra-indications (stabilizing hardware is typically MRI safe) could enroll but would not receive an MRI.

### 2.4 Randomization

Participants were randomized via computer-generated allocation to 8 weeks of in-person CMR with a therapist or in-person adaptive fitness at the Courage Kenny Rehabilitation Institute (45 min, 3x/week). Raters, the biostatistician, brain technicians and brain imaging analysts were blinded to group allocation.

A CMR-certified teacher delivered all CMR sessions in Van de Winckel’s wheelchair-accessible Brain Body Mind Lab. The sessions were video-recorded and logged. For adaptive fitness, a physiatrist assessed participants post-baseline in the Brain Body Mind Lab to confirm safety and provided written physical authorization (as was required by the adaptive fitness center). Training adherence was monitored via log sheet at the Courage Kenny Rehabilitation Institutes. Both interventions were classified as minimal risk: CMR due to low-intensity movements and kinesthetic imagery; and adaptive fitness as it is standard care available in the community for adults with SCI.

### 2.5 Study procedures

Neuropathic pain was assessed with the Numeric Pain Rating Scale (NPRS), and sensorimotor function was evaluated with the ASIA Impairment Scale (AIS), also called, International Standards for Neurological Classification of Spinal Cord Injury (ISNCSCI), and Neuromuscular Recovery Scale (NRS). All clinical and behavioral assessments (questionnaires) were conducted at baseline, post-intervention, and three-month follow-up. Functional magnetic resonance imaging (fMRI) included resting-state and 4 tasks: 1– feeling the left leg; 2–imagining the feeling of moving the left leg, 3-whole-body movement imagery, and 4-a sensation task. MRI testing was performed at baseline and after post-intervention.

CMR sessions and AIS(27) exams were conducted in Van de Winckel’s Brain Body Mind Lab at the University of Minnesota (UMN). MRI scanning took place at the UMN’s Center for Magnetic Resonance Research (CMRR), with participants screened via a CMRR-prescreening questionnaire. Participants completed behavioral questionnaires on REDCap with trained study staff via secure UMN Zoom.

### 2.6 Aims

Our first aim was to determine whether 8 weeks of CMR or adaptive fitness improved sensory and motor function in adults with SCI, with CMR hypothesized to show greater improvement on performance-based clinical tests of sensorimotor function. Secondary outcomes included neuropathic pain intensity, spasm, mood, self-reported function, body awareness, self-efficacy, sleep, and SCI-related quality of life (namely, positive affect, resilience, self-esteem, ability to participate, pain behavior, and independence), all hypothesized to improve after CMR.

Our second aim was to determine whether 8 weeks of CMR or adaptive fitness increased brain activity and connectivity in MBR– and sensorimotor brain areas and networks in adults with SCI, assessed by resting-state and 4 task-based fMRI. We hypothesized that CMR would increase parietal operculum, insula, and posterior cingulate cortex connectivity with other sensorimotor, body awareness, and visuospatial-related brain areas, concurrent with clinical sensorimotor improvements.

### 2.7 Interventions

#### 2.7.1 Cognitive Multisensory Rehabilitation

“Cognitive Multisensory Rehabilitation” (CMR) is a physical therapy approach from Italy that focuses on restoring MBR to improve actions and the way the body is interacting with the environment. Guided by the CMR-certified therapist, participants use body signals, sensations, and kinesthetic imagery to solve multisensory discrimination exercises that require participants, for example, differentiating various positions of the limbs in space, or recognizing different limb positions relative to other body parts. Through these exercises, participants also reintegrate correct touch and pressure sensation because the exercises can include the use of different textures or sponges with varying resistance that can be placed on or under different parts of the body according to the specific exercise. The multisensory discrimination exercises are used as a means to recover MBR but the key to this MBR restoration is the guidance that the therapist provides, for example, through left-right comparisons, contrasting past memories and present situations, body scans, comparison of their position or movements through demonstrations by the therapist or through drawings. While the CMR delivery is standardized, exercise choice and complexity as well as body parts addressed is personalized to each patient.

CMR addresses body part relationships such as (i) leg position in space and relative to the pelvis and the upper body (ii) the dimensions and length of the legs, the dimension of the pelvis, and sensation of the pelvis as a central body reference. (iii) the relationship between the left and right side of the body, and the relationship between the pelvis and the feet. Further, improvements in touch and pressure sensation are obtained through the experience of the exercise. By reinvolving these body parts in MBR, multisensory signals become consistently perceived and integrated. As sensory feedback is essential for movement, sensory gains create the conditions for motor recovery. Once the brain accurately maps the body, neuropathic pain and spasms are also reduced or resolved. Concrete examples of CMR exercises are provided in supplementary **Data Sheet 1**. Further details can be found in the protocol paper and our prior publication of CMR in adults with SCI-related neuropathic pain.(10,16)

#### 2.7.2 Adaptive fitness

The active control intervention was an adaptive fitness program, reflecting standard, community-accessible exercise available to adults with SCI post-rehabilitation. Participants used state-of-the-art adaptive gym equipment at the Courage Kenny Rehabilitation Institutes in Golden Valley and Stillwater locations in Minnesota.

The fitness centers were specifically designed for accessibility, with equipment spaced for movement, transfers, and (if needed) caregiver assistance, and several machines accessible from a wheelchair. Equipment included treadmills, ellipticals, NuSteps, SciFit upper body ergometers, SkiErgs, rowing machines, upright and recumbent stationary bicycles, chest presses, rear rows, leg extensions, leg curls, leg presses, uppertones, free weights, and kettlebells. Bodyweight-supported treadmill training was excluded to ensure the program reflected standard exercises that participants could complete without the need for highly specialized robotic or harness equipment.

Prior to starting the intervention, participants completed an evaluation session with a trained fitness specialist at the Courage Kenny Rehabilitation Center. The specialist then designed a personalized adaptive fitness program tailored to their needs and preferences. Participants then performed their customized program under the supervision of a trained fitness specialist for 45min/session, 3x/week.

The 45 min workouts differed per person but could be, for example, “25-30 min on the Nustep + 15-20min leg press”; “cardio exercises + treadmill + chest and shoulder exercises”; “chest press + shoulder press + chest and back press + strings and free weights”; “stretching on the mat + legs, shoulders and biceps strengthening”.

### 2.8 Primary clinical outcomes

AIS (28,29) clinical exam identifies the extent and severity of SCI, and takes about 40 min to administer. It includes a pinprick test (sharp versus dull with a safety pin); a light touch test (with a cotton ball) and muscle strength testing, performed by a trained clinician or therapist. These sensation tests were scored from 0-54 per side (anus test excluded), and muscle strength for upper and lower limbs were scored from 0-25 per limb per side. Higher scores indicate better function. The AIS was assessed at baseline, post-intervention, and at the 3-month follow-up.

The NRS (29,30) includes 11 functional tasks focused on trunk balance, and activities with the trunk (core), arm and hand function, and lower extremity tasks (e.g., sit-to-stand, weight-bearing, taking steps, and stepping over an obstacle). It measures non-compensatory functional recovery with higher scores indicating better performance. NRS administration requires online training and certification and takes about 40 min to administer.

### 2.9 Secondary clinical outcomes

The Numeric Pain Rating Scale (NPRS) (31) was used to assess highest, average (i.e., most of the time), and lowest neuropathic pain intensity in the prior week, rated on a scale with scores ranging from 0 (no pain) to 10 (worst pain ever).

The Douleur Neuropathique 4 (DN4) is a diagnostic tool used to identify and assess neuropathic pain.(32)

For the Patient-Specific Functional Scale (PSFS), participants chose three functional activities that were important to them, for which progress was measurable, and that they were unable to do or complete at baseline because of their neuropathic pain. Participants rated them between 0 (unable to do the activity) and 10 (able to do the activity).(33)

The Spinal Cord Injury Functional Index/Assistive Technology (SCI-FI/AT) consists of 32 self-report items, rated by self-reported level of difficulty in doing daily activities with assistive devices if needed but not with the help of another human (i.e., 0-“unable to do” to 4-“no difficulty”). The scale reliably measures function in the domains of basic mobility, self-care, fine motor function, and ambulation. Test administration takes about 10 min.(34,35)

Muscle spasms were rated for frequency and severity with the Penn Spasm Frequency Scale.(36)

The Pittsburgh Sleep Quality Index (PSQI) consists of 7 components with total scores ranging from 0-21, where lower scores indicate less difficulty in the assessed areas, and a score >5 indicating poor sleep quality.(37)

The Spielberger State Trait Anxiety Inventory (SAI and TAI) assesses trait anxiety (general feelings) and state anxiety (present feelings). Both scales are scored from 20-80; lower scores indicate less anxiety.(38)

The Patient Health Questionnaire (PHQ-9) consists of 9 items scoring symptoms of depression. Each item is scored from 0-3, with total scores ranging from 0-27; higher scores indicate greater symptoms of depression.(39)

The Tampa Scale for Kinesiophobia (TSK) consists of 17 items scored from 1-4 (total 17-68) with lower scores indicating greater fear of re-injury.(40)

With the Autonomic Standards Assessment form (ANS), participants self-reported on cardiovascular health, bladder, bowel and sexual function, the latter three scored as normal, reduced, or absent.(41)

The Moorong Self-Efficacy Scale (MSES) consists of 16 items each scored from 1-7, with total global scores ranging from 16-112. The MSES assesses self-efficacy related to everyday life activities and is designed specifically for people with SCI. Higher scores indicate greater self-efficacy.(42,43)

The Functionality Appreciation Scale (FAS) assesses with 7 items how appreciative a person is for their body functionality.(44,45)

The Postural Awareness Scale (PAS) is a 12-item scale, assessing self-reported awareness of body posture.(46)

The Revised Body Awareness Rating Questionnaire (BARQ-R) is a 12-item scale assessing body states, e.g., how tension in the body affects one’s body awareness and function in daily life. Each item is scored from 0-3, with total scores ranging from 0-36. Lower scores indicate better body awareness.(47,48)

The Multidimensional Assessment Interoceptive Awareness – version 2 (MAIA-2) (37 items) measures 8 interoception domains through self-report: noticing (awareness of uncomfortable, comfortable, and neutral body sensations); not-distracting (tendency to not ignore or distract oneself from sensations of pain or discomfort); not-worrying (tendency not to worry or experience emotional distress with sensations of pain or discomfort); attention regulation (ability to sustain and control attention to body sensations); emotional awareness (awareness of the connection between body sensations and emotional states); self-regulation (ability to regulate distress by attention to body sensations); body listening (active listening to the body for insight), and trusting (experience of one’s body as safe and trustworthy).(49)

The Kinesthetic and Visual Imagery Questionnaire (KVIQ) assesses the visual imagery clarity and kinesthetic imagery intensity of sensations from a first-person perspective (i.e., seeing/observing someone do it or imagining how it would feel to do it oneself). Each item is scored 1-5 and summed for a global score, higher scores indicating better imagery.(50)

The Spinal Cord Injury Quality of Life measurement system (SCI-QOL) encompasses 22 subdomains across 4 broad domains of physical-medical health, emotional health, social participation, and physical functioning. We used 6 short-form subscales: positive affect (10 items), resilience (8 items), self-esteem (8 items), ability to participate (10 items), independence (8 items), pain behavior (7 items).(51)

The Craig Handicap Assessment and Reporting Technique-Short Form (CHART-SF) assesses the degree by which a person with SCI remains with an impairment or disability. It measures the type and level of assistance needed physically and cognitively; mobility reflected by the level of physical activity and transportation needs; occupation (how time is spent); social interactions; and financial resources (economic self-sufficiency). Each of six domains is scored from 0-100, with total scores ranging from 0-600. Higher scores indicate less impairment.(52)

Participants attended weekly phone or Zoom check-ins with a research assistant, blinded to group allocation, who recorded any adverse events (also unrelated to the study), use of over-the-counter or prescribed medications for spasms (or for neuropathic pain, if present), recent illnesses, healthcare utilization and/or recent hospitalizations, and information related to community integration. The CMR therapist and staff at Courage Kenny recorded adherence. They also provided information about the content of the intervention performed that week. Another research staff member, not blinded to the intervention, asked about perceived effects of the intervention and content of the interventions. We allowed concomitant medication prescribed or recommended by their clinical care team throughout the trial, as well as other treatments/therapies related to their standard of care such as stretching. However, participants did not receive additional balance, gait, or strengthening exercise outside of the study interventions, or any additional pain management treatments aside from prescribed or over the counter medication.

### 2.10 Primary brain imaging outcomes

Brain imaging was performed on a Siemens 3-T Prisma scanner at CMRR. Participants were scanned for two hours. Full acquisition details are reported in a previous protocol paper.(10) Briefly, structural MRI acquisition included T1-weighted magnetization-prepared rapid acquisition with gradient echo (MPRAGE) [repetition time (TR)=2.5s; echo time (TE)=4.5ms; 0.8mm isotropic voxels], and T2-weighted sampling perfection with application-optimized contrasts using different flip angle evolution (SPACE) [TR=3.2s; TE=565ms; 0.8mm isotropic voxels].

Resting-state and task fMRI scans were acquired with T2*-weighted multiband echo planar acquisition tilted 30 degrees relative to the anterior commissure–posterior commissure (AC-PC) plane according to auto-align software, enabling whole-head blood-oxygen-level-dependent (BOLD)–contrast with optimal temporal and spatial resolution and to reduce signal dropout [TR = 0.8s; TE = 37ms; flip angle = 55degrees; 72 slices; multiband factor 8; 2mm isotropic voxels]. The resting-state fMRI imagery lasted 12min 10sec. Participants were asked to maintain eye fixation with a restful mind.

Resting-state connectivity and fMRI tasks characterized brain areas and networks involved in sensorimotor and MBR function in adults with SCI. Seeds in the parietal operculum, insula, and posterior cingulate cortex were selected based on our CMR work in stroke and SCI, as well as other research demonstrating their importance in MBR.(8–11)

#### Task 1: Mental body scan (8 min 15 sec)

Participants focused with gentle, non-judgmental awareness on sensations of the left leg (having reduced sensation and movement). In other words, they perform a gentle focused mental body scan and register any feelings of sensation in that limb. We alternated this task with rest periods. This task was selected based on our prior work in adults with chronic low back pain showing parietal operculum and posterior parietal cortex activation after body awareness training (Qigong),(53) and evidence from other brain imaging research that mindful body scanning increases insula connectivity.(54)

#### Task 2: Kinesthetic imagery of moving the limb (7 min 58 sec)

Participants were asked to imagine the feeling of moving the left leg without actually moving (i.e., gentle non-judgmental awareness of any sensations during an imagined movement), alternated with rest. We chose this task based on tasks with mindfulness meditation showing insula and parietal operculum activation.(55)

#### Task 3: Whole-body movement task (19 min 51 sec)

After viewing a video demonstration, participants performed a kinesthetic imagery of a whole-body movement (a Qigong movement)(56) without moving. They imagined the feeling of being upright, hands moving symmetrically toward and away from the body in a horizontal line (at the level of the navel) with guided breathing, while imagining the legs bending and straightening. This task was selected based on studies showing substantial spatial overlap of areas activated during motor imagery vs. motor execution.(57–60) Our prior studies showed parietal operculum and posterior parietal cortex activation during this task in adults with chronic low back pain.(53)

#### Task 4: Sensory stimulation task (18 min 32 sec)

The pads of the big toes and thumbs were gently stroked with a towel without participants’ knowledge of when stroking occurred or where on the hand or feet it would occur. After the scan, participants were asked whether and where they felt stimulation. We chose this because in our previous CMR study in adults with SCI-related neuropathic pain, greater activation was found after CMR in the bilateral postcentral gyrus (sensory function), angular and supramarginal gyri, superior parietal lobe (part of the posterior parietal cortex, related to visuospatial body maps); left insula (pain processing, body awareness), and frontal operculum.(16)

### 2.11 Statistical analysis

#### 2.11.1 Clinical outcome analysis

##### Sample size calculation

Based on our prior pilot study,(16) baseline to post-CMR average differences were 8.81±5.37 points for the pin prick test, 7.50±4.89 for the touch test, and 3.87±2.80 for lower limb motor function. Assuming similar effect CMR effects and no change after adaptive fitness, the sample size of 8 participants per group provides 89%, 85%, and 76% power to detect significant group differences for pin prick, touch, and lower limb motor function, respectively, at an overall two-sided significance level of α=0.05 with Bonferroni-correction for the three primary endpoints. The present data are essential to determine effect sizes to power a future larger randomized controlled trial.

We summarized all quantitative variables with descriptive statistics at each time point per group. We used R for all statistical analyses. Given the small sample size, we reported Cohen’s d effect sizes per group for pre-post changes and pre-3-month follow-up changes in sensorimotor function measures and other clinical outcomes.(61)

A UMN Clinical and Translational Science Institute data integrity monitor and quality assurance reviewer monitored the study every 6 months.

#### 2.11.2 Brain imaging analysis

Preprocessing was done through the Human Connectome Project preprocessing pipeline. We used the CONN functional connectivity toolbox to process fMRI data with established standardized controls for multiple comparisons (Statistical Parametrical Mapping family-wise error correction methods).(62) Data underwent realignment, scrubbing, artifact detection, and CompCor denoising.

For the primary analyses, we used voxel-wise whole-brain seed-based functional connectivity analysis of resting-state data using Pearson correlation coefficients, focused on parietal operculum, insula, and posterior cingulate cortex regions-of-interest for resting-state connectivity and exploratory multivariate pattern analyses. Task-based fMRI underwent the same preprocessing pipeline. We estimated voxel-wise brain activation for each participant using a general linear model.

Within-group pre-post changes in fMRI task-based brain activation and resting-state connectivity (with Fisher’s Z transformation) were tested using paired t-tests or Wilcoxon signed-rank tests.

We tested the effect of CMR on brain imaging outcomes using mixed-effects models implemented in FSL. Cluster-based correction for multiple comparisons was performed using a cluster-forming threshold of Z > 2.3 and a cluster-level significance threshold of p < 0.05 to control the family-wise Type I error rate.

## 3 Results

**Fig. 1** displays the CONSORT flow diagram. Of 27 recruited participants with SCI, 3 were excluded (baclofen pump, cognitive impairment from brain injury, inability to self-transfer), and 8 withdrew during the study for various reasons [e.g., illnesses unrelated to the study (n=2), i.e., acute ulcer wound, COVID-19; the baseline fitness level of participants was too high to participate in the study (n=3); or were not responding to communication after consenting (n=3)]. We collected MRI scans for 13 participants at baseline. Reasons for not having an MRI were either a bullet in the body or a heating risk of a Songer cable as part of a cervical fusion.

**Fig. 1.**
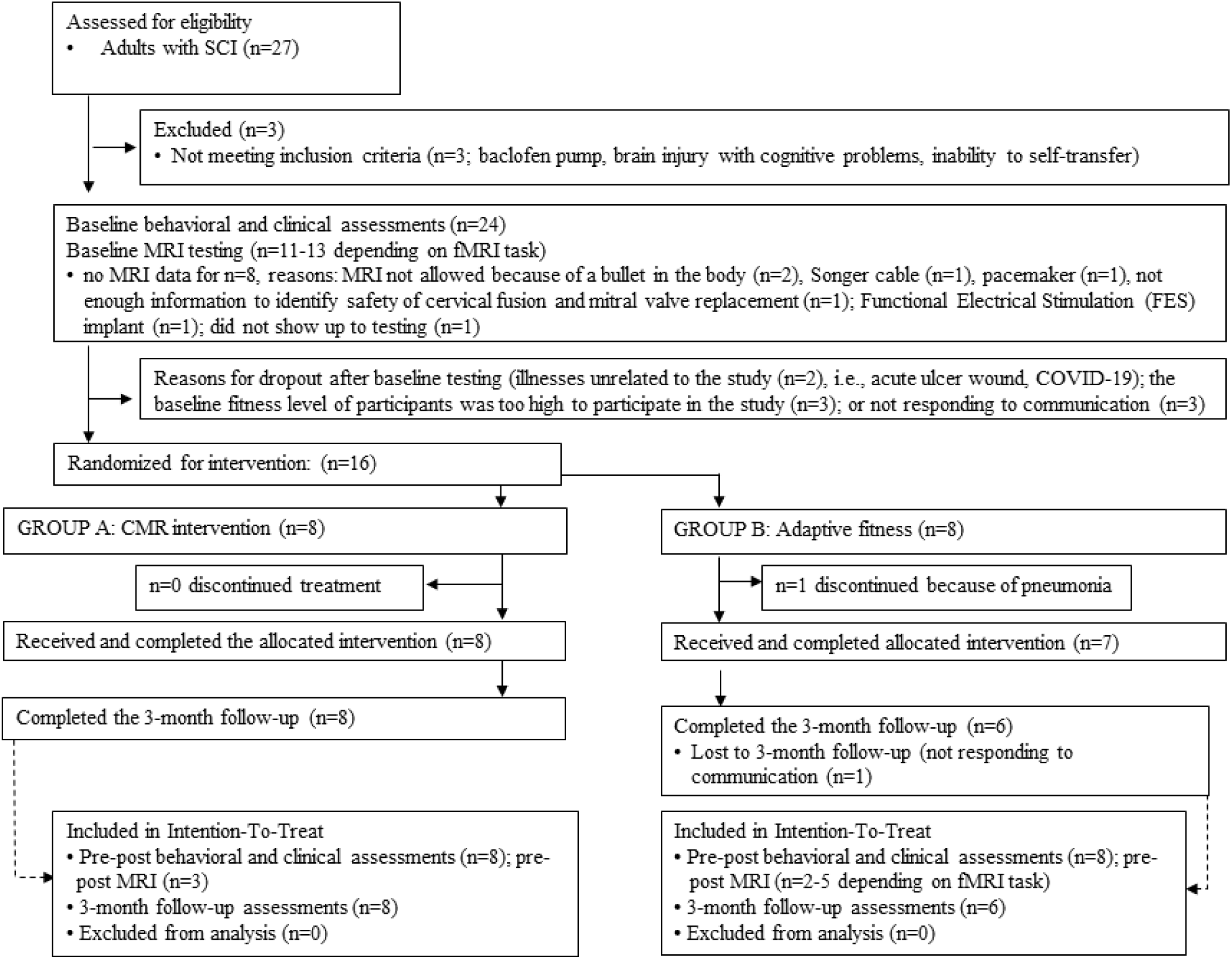
CONSORT flow diagram.

Sixteen adults were randomized to the interventions. The group had an average age of 52 ± 8 years (range of 38-64 years), 6 women, and 12 men; 25% African American, 75% White non-Hispanic adults, and 19% living in rural areas. They were 13 ± 10 years after SCI (range of 1-29 years). Six adults had tetraplegia, one had Brown-Sequard, and 10 had paraplegia. There were no significant baseline differences in AIS or NRS outcomes between the groups. The demographic and clinical information of the 16 participants are detailed in **Table 1**.

All 8 participants completed the *CMR intervention* and all 8 completed the 3-month follow-up phase. In the fitness group, 8 participants started the 8-week *adapted fitness intervention* and one person withdrew during the intervention phase because of pneumonia. Of the remaining 7 that completed the 8-week intervention, 6 participants completed the 3-month follow-up phase (1 did not respond to communication at the time of the follow-up testing). All participants averaged 135 min/week (3 sessions of 45min/week) of intervention throughout the 8 weeks. On occasion, missed sessions due to illness were made up the following week. The final analysis included 15 participants in total: 8 participants in the CMR group and 7 people in the fitness group, with n=6 for the 3-month follow-up in the fitness group.

### 3.1 Clinical outcomes

Clinical outcomes for Aim 1 are reported in **Table 2** with effect sizes for post-intervention and 3-month follow-up vs baseline, reflecting the dual burden of SCI across sensorimotor function and neuropathic pain.

CMR yielded greater sensorimotor improvements than adaptive fitness post-intervention. AIS effect sizes were large for CMR (touch: Cohen’s d=1.54; pinprick: d=1.83; lower limb motor function: d=1.32) and small-to-moderate for adaptive fitness (touch: d=0.49; pinprick: d=0.53; lower limb motor function: d=0.74, **Fig. 2**). NRS effect sizes were also larger for CMR (core: d=2.19; upper limb: d=0.69; lower limb: d=0.74) than adaptive fitness (core: d=0.73; upper limb: d=0.34; lower limb: d=0.00, **Fig. 3**).

**Fig. 2.**
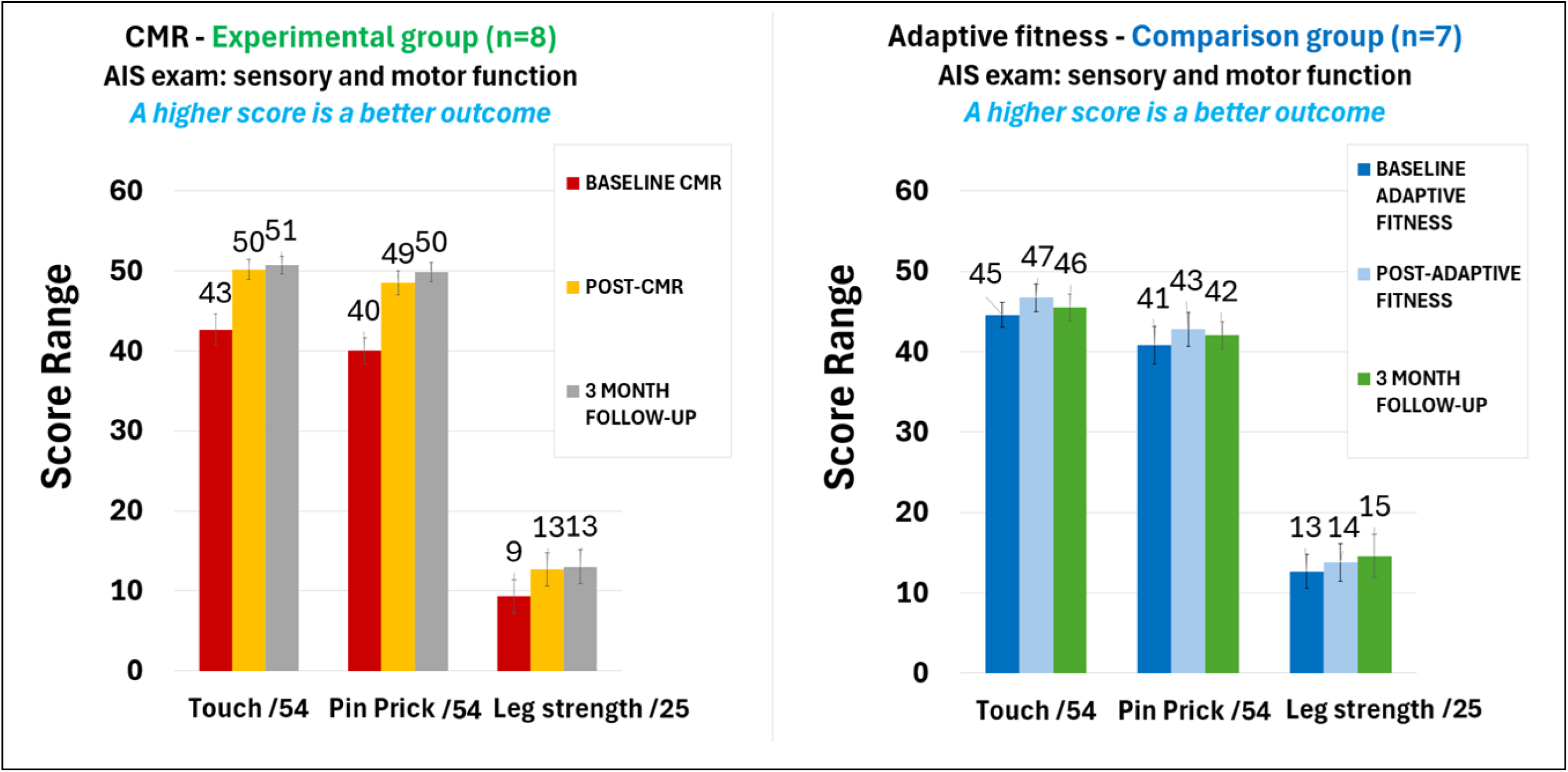
AIS exam. Bar graphs show average left and right scores for touch, pinprick sensation, and lower limb strength at baseline, post-intervention, and 3-month follow-up for the CMR and adaptive fitness groups.

**Fig. 3.**
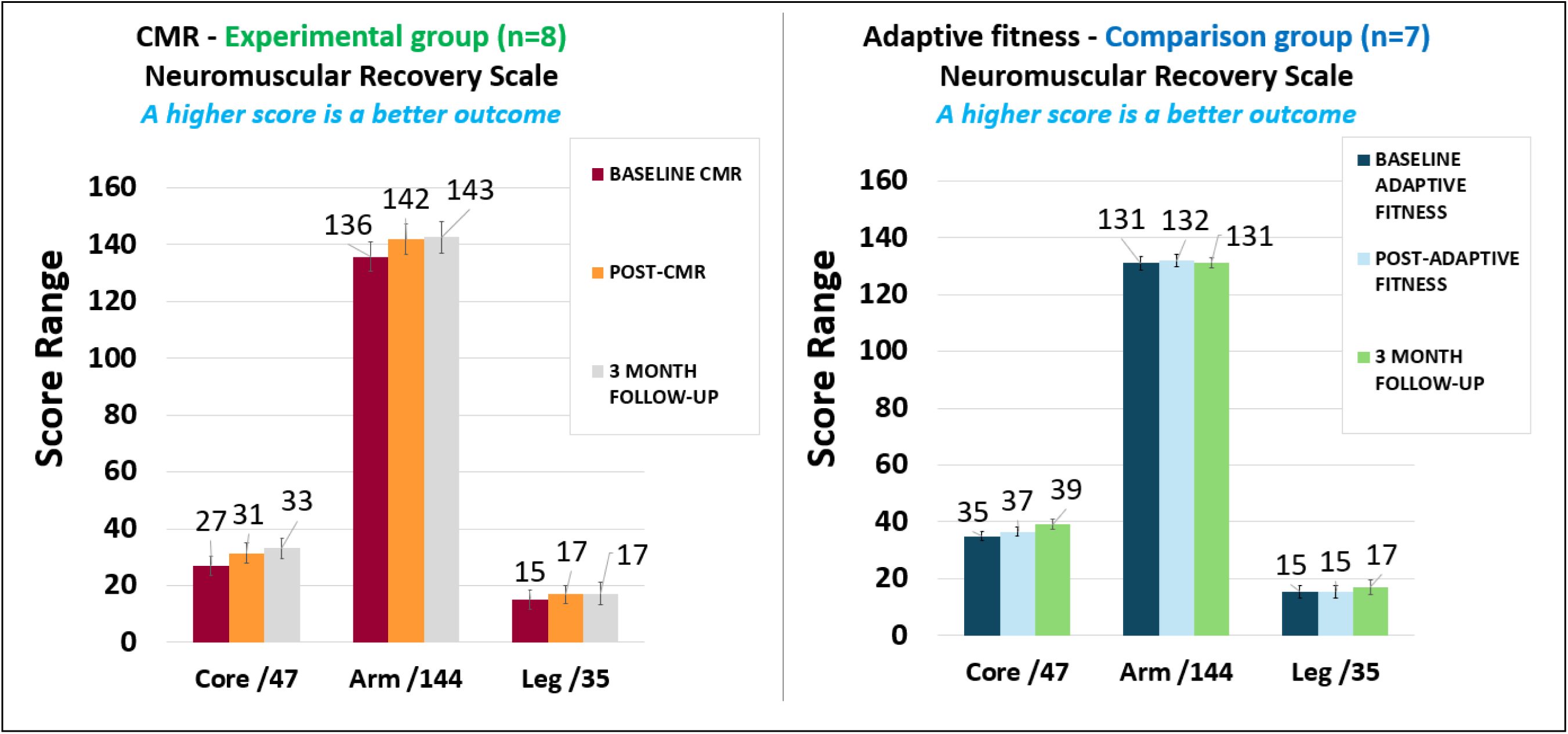
NRS exam. Bar graphs show scores for core strength, upper limb and lower limb function at baseline, post-intervention, and 3-month follow-up for the CMR and adaptive fitness groups.

At 3-month follow-up compared to baseline, CMR maintained large AIS effect sizes (touch: d=1.73; pinprick d=2.07; lower limb motor function: d=1.28, **Fig. 2**) and moderate-to-large NRS effect sizes (core: d=1.74; upper limb: d=0.79; lower limb: d=0.78). Adaptive fitness group showed moderate AIS effect sizes (touch: d=0.56; pinprick d=0.74; lower limb motor function: d=0.63) and moderate-to-large NRS effect sizes (core: d=0.62; upper limb: d=0.85; lower limb: d=0.65, **Fig. 3**).

Notably, 53% of participants across both groups (n=4 in each group) achieved maximum upper limb NRS scores at baseline, reflecting 7 individuals with paraplegia, and 1 person with quadriplegia. The person with quadriplegia had a dislocated rather than a fractured spine, which may have contributed to a complete motor recovery, yet the participant still had moderate pinprick sensory impairment throughout the trunk at baseline. In sum, this expected ceiling effect constrained the observable upper limb change scores across the cohort.

Neuropathic pain results across the full cohort (n=15), showed a tendency toward divergent outcomes between the two interventions. However, interpretation is limited by the small sample size, given that only 10 out 15 participants had neuropathic pain at baseline, and with large NPRS standard deviations, reflecting substantial inter-individual variability (baseline highest neuropathic pain severity range 3-10/10).

Accounting for this variability and the fact that 5 participants did not have pain at baseline (3 in the CMR group, 2 in the adaptive fitness group), CMR showed small post-intervention effect sizes for pain reduction (highest NPRS: d=0.48; average NPRS: d=0.46) and small-to-moderate effects at 3-month follow-up, compared to baseline (highest NPRS: d=0.63; average NPRS: d=0.31). CMR also reduced severity of neuropathic pain characteristics (DN4) with a moderate effect size sustained at follow-up (d=0.59 at both timepoints).

The adaptive fitness group showed a tendency toward increased neuropathic pain post-intervention (highest NRPS, d=-0.19) and neuropathic pain characteristics (DN4, d=-0.38), but a slight decrease in average pain (NPRS, d=0.38). At 3-month follow-up compared to baseline, highest NPRS was unchanged (d=0.00), average NPRS slightly decreased (d=0.41), neuropathic pain characteristics slightly worsened in severity (d=-0.41).

In the CMR group, 2 participants took neuropathic pain medication. One reduced Lyrica by 88% at the 3-month follow-up, along with 2-point reduction in highest neuropathic pain. The other participant reduced medical cannabis by 50% while maintaining the Gabapentin dosage, with no change in pain intensity levels. All medication dosage changes were authorized by their treating physicians. In the fitness group, 2 out of the 4 participants on pain medication increased Gabapentin by 50-66% (one reporting more tingling, the other a 2-point reduction in highest pain at follow-up). Another person reduced Hydrocodone by 29% post-intervention but increased it by 12.5% at follow-up (compared to the baseline dosage), while keeping Amitriptyline stable and pain slightly worsening by one point. The last participant maintained stable Gabapentin and Baclofen dosages across the study with no pain changes.

Beyond our sensorimotor and neuropathic pain indices, we evaluated a comprehensive suite of secondary clinical measures to capture the broader functional and psychosocial impacts of the interventions. These outcomes were analyzed across the full cohort (n=15) except for SCI-FI/AT, which has a separate scoring system for paraplegia and quadriplegia.

#### 3.1.1 Function

Both groups showed large PSFS effect sizes, for self-identified functional goal attainment (CMR: d=1.99, adaptive fitness: d=1.07), sustained at follow-up (CMR: d=1.95; adaptive fitness: d=1.81).

For SCI-FI/AT paraplegia, CMR (n=5) showed large effect sizes for basic mobility (d=1.47), self-care (d=1.41), and fine motor function (d=1.38), with moderate improvements in ambulation (d=0.61), all maintained at follow-up (d=1.37; d=1.41; d=1.27; d=0.61, respectively). The adaptive fitness results (n=4 at post-intervention; n=3 at follow-up) were more varied and should be interpreted cautiously given the small sample size. Basic mobility improved moderately post-intervention (d=0.78) but decreased to baseline levels at follow-up. Self-care declined after the fitness intervention, then improved at follow-up (d=0.58). Fine motor function was unchanged after fitness (d=0.00) but improved at follow-up (d=0.58). Finally, ambulation improved post-intervention (d=0.50) and further at follow-up (d=1.09).

For adults with quadriplegia (n=3 per group), CMR showed large improvements in SCI-FI/AT basic mobility (d=1.16) and self-care (d=1.16) at both timepoints, versus moderate-to-large fitness improvements (selfcare: d= 0.58; basic mobility: d=0.87). All 3 CMR participants had maximum fine motor scores at baseline (d=0.00) precluding improvement. Fitness showed large fine motor function gains (d=1.00) at both timepoints. Ambulation improved in both groups, with moderate CMR effect sizes (d=0.58) and large fitness effect sizes (d=1.12) at both timepoints.

Mobility, as indexed by the CHART-S, reflected gains following CMR (d=1.18) compared to adaptive fitness (d=0.37), with moderate effect sizes in both groups at the 3-month follow-up (d=0.76 for CMR; d=0.55 for fitness).

#### 3.1.2 Body awareness and motor imagery

KVIQ showed large CMR effect sizes at both timepoints (d=1.02; d=1.07) while fitness showed moderate improvements post-intervention (d=0.73) but a decline at follow-up (d=-0.60).

CMR produced immediate and sustained FAS improvements (d=0.61; d=0.52), whereas fitness had minimal impact (d=-0.13; d=0.10, respectively). PAS improved moderately in both groups post-intervention (CMR: d=0.60; fitness: d=0.54) but there were only small improvements compared to baseline at follow-up (CMR: d=0.25; fitness: d=0.34).

BARQ-R (body tension, emotional body awareness, awareness of breathing) showed no immediate post-intervention effects, but there were moderate CMR improvements (d=0.59) and a small fitness improvement (d=0.43) at follow-up.

Pertaining to interoception, the eight domains of MAIA-2 showed small improvements immediately post-CMR (ranging from d=0.11 to 0.43), except for attention regulation (d=-0.42), growing to small-to-large improvements at follow-up (ranges of d=0.11 to 1.14; except for attention regulation: d=-0.12). Fitness only improved in 3 subdomains with small-to-moderate effect sizes (d=0.25 to 0.77), while 5 subdomains worsened (d=-0.38 to –1.37). This pattern persisted at follow-up (3 improved: d=0.04-0.15; 5 subdomains worsened compared to baseline levels: d=-0.24 to –0.66).

#### 3.1.3 Spasticity, autonomic function, sleep, and psychosocial wellbeing

Spasticity frequency and intensity improved comparably in both groups post-intervention and at follow-up (ranges d=0.38 to 0.65) except for spasm frequency in the CMR group at follow-up, which was unchanged compared to baseline (d=0.00).

Autonomic function improved moderately with CMR at both timepoints (d=0.68) while fitness showed slight worsening post-intervention that nearly resolved to baseline at follow-up (d=-0.55; d=-0.18).

Both groups also experienced small improvements in sleep quality (PSQI; CMR: d=0.34 and 0.12; fitness: d=0.31 and 0.22, respectively post-intervention and at 3-month follow-up).

Psychosocial wellbeing outcomes presented a more nuanced profile. Trait anxiety decreased substantially more with CMR (d=1.53; d=1.59) than fitness (d=0.56; d=0.72) at both timepoints. TSK improved in both groups at 3 months follow-up (CMR: d=0.75, fitness: d=0.78), but state anxiety, PHQ-9, and MSES showed no meaningful changes in either group.

### 3.2 Brain imaging outcomes

For aim 2, baseline results confirmed the validity of our tasks and region-of-interest selections. Resting state seeds (insula, parietal operculum, and post-cingulate cortex) connected with sensorimotor areas (pre-and postcentral gyri), pain perception and body awareness regions (parietal operculum and insula), and visuospatial body map areas (superior parietal lobe, supramarginal gyrus, and angular gyrus). Task-based fMRI similarly showed baseline activation in these same brain regions.

Concurrent with clinical improvements, resting-state fMRI showed increased brain connectivity after CMR compared to baseline (**Fig. 4**, **Table 3**), specifically, stronger connectivity between the right parietal operculum (as region-of-interest) and the right precentral gyrus (**Fig. 4A**), and between the right posterior cingulate cortex (as region-of-interest) and the left precentral gyrus, left insula, and left parietal operculum (parts OP1/OP4, (**Fig. 4B**). No connectivity changes were found in the fitness group.

**Fig. 4.**
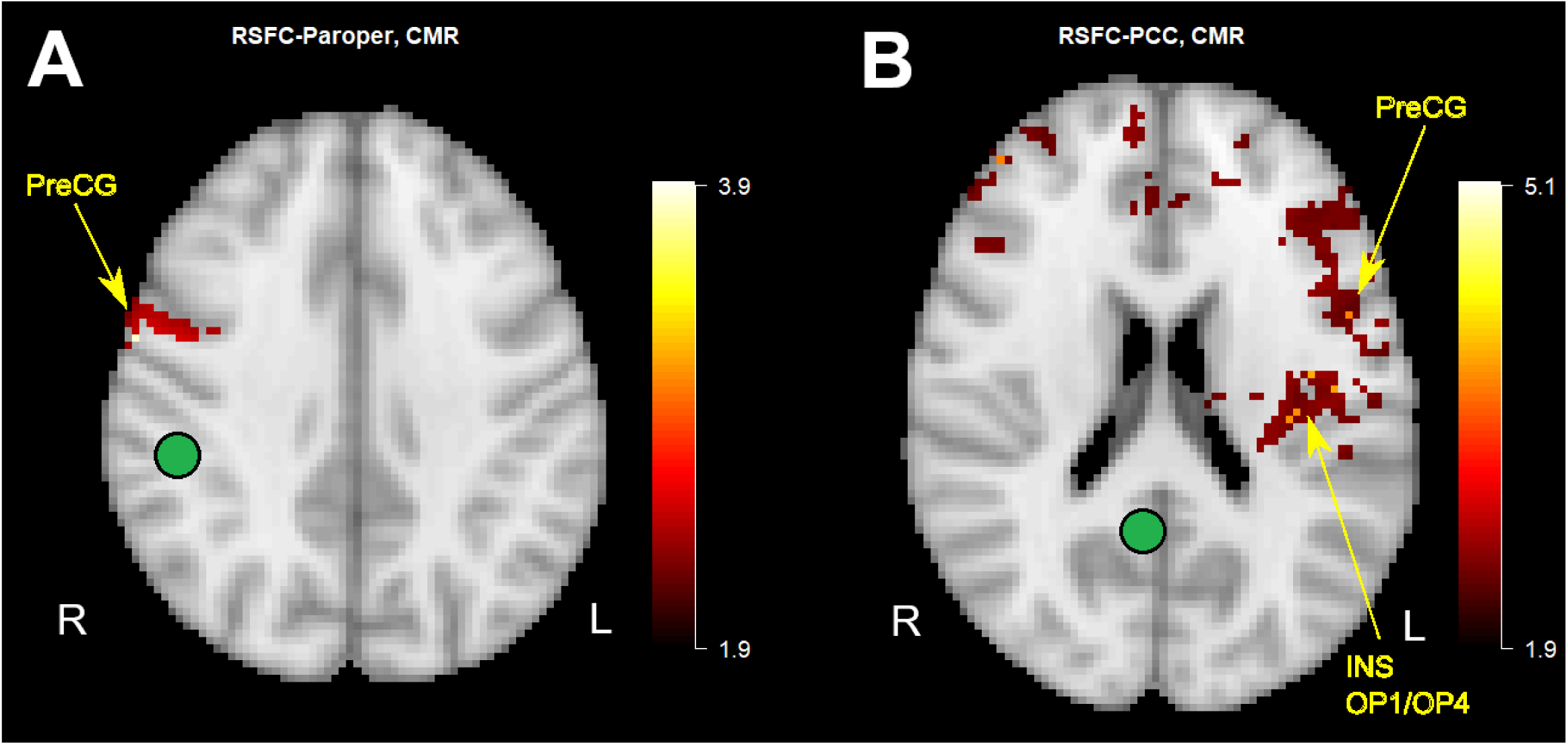
Pre-post CMR changes in connectivity strength. (**A**) Stronger parietal operculum network connectivity after 8 weeks of CMR: right parietal operculum (ROI) to right precentral gyrus (PreCG). (**B**) Stronger posterior cingulate cortex network connectivity after CMR: right posterior cingulate cortex (ROI) to left precentral gyrus (PreCG), left insula (INS), and left parietal operculum (OP1/OP4).

CMR also increased task-based brain activity compared to baseline: Imagining *feeling the left leg*, activated the pre– and postcentral gyri, and superior parietal lobe to a greater extent post-CMR vs baseline (**Fig. 5**, **Table 3**). The *whole-body imagery task* produced greater activation post-CMR in the right pre– and postcentral gyrus, bilateral supramarginal gyrus, superior parietal lobe, and angular gyrus (**Fig. 6A**, **Table 3**).

**Fig. 5.**
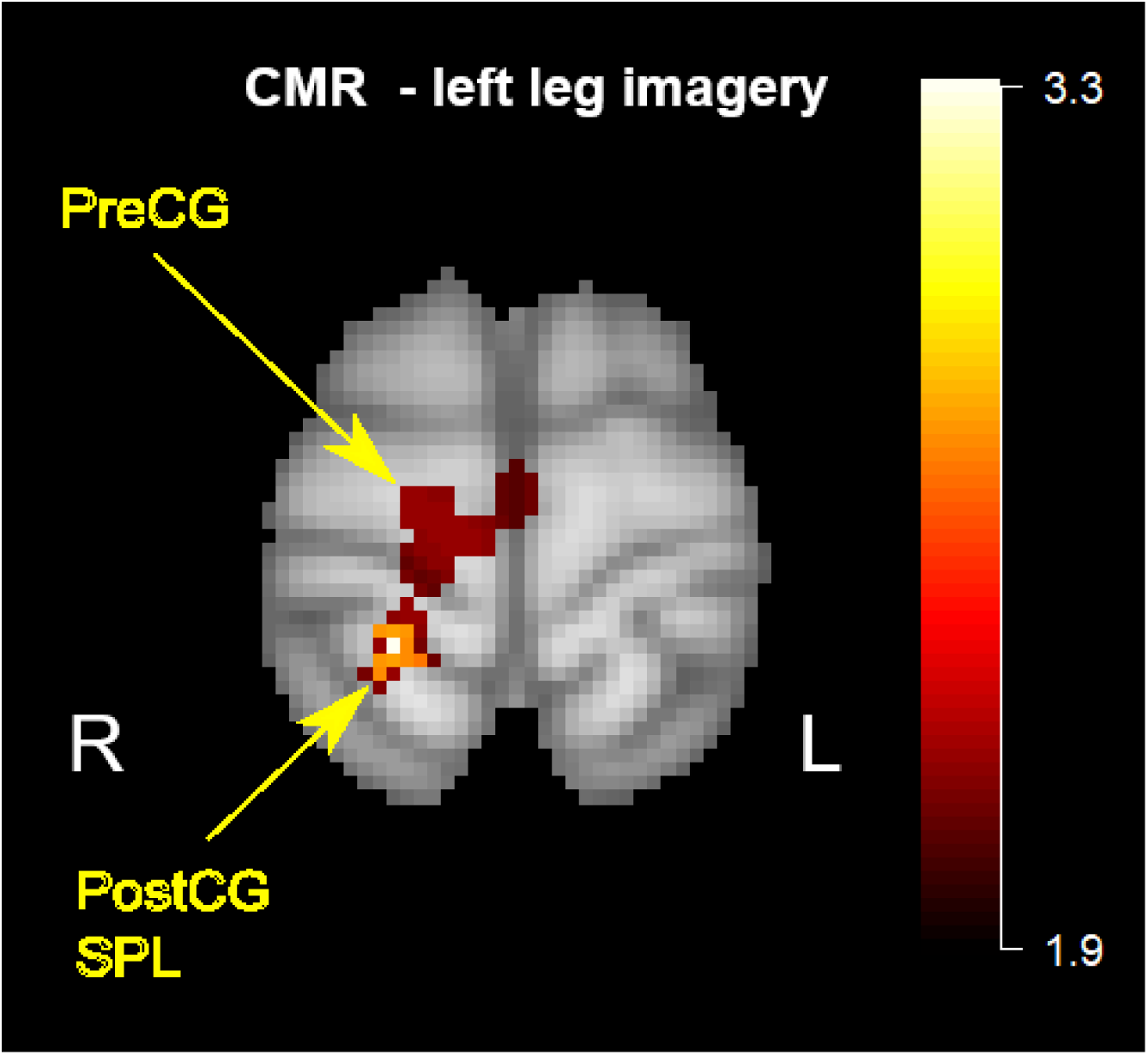
Pre-post CMR changes in brain activity during leg kinesthetic imagery. Post-CMR, enhanced brain activity was observed in the right pre– and postcentral gyri, and right superior parietal lobe.

**Fig. 6.**
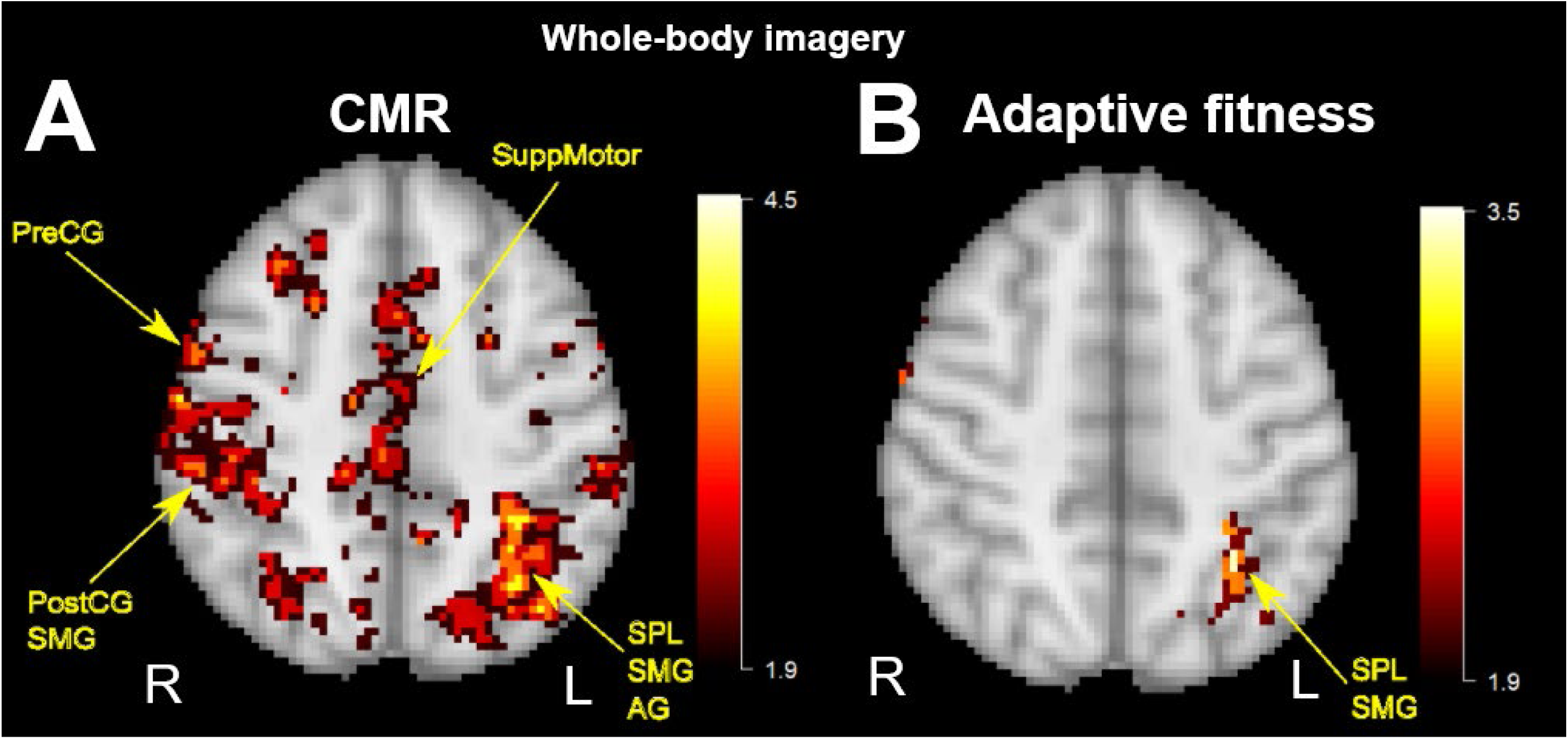
Pre-post changes in brain activity during whole-body imagery for CMR and adaptive fitness. (**A**) Post-CMR increased activation in the supplementary motor area (SuppMotor), right pre-and postcentral gyrus (PreCG, PostCG), bilateral supramarginal gyrus (SMG), superior parietal lobe (SPL), and angular gyrus (AG). (**B**) Post-adaptive fitness, only smaller areas of the left supramarginal gyrus (SMG) and superior parietal lobe (SPL) were activated.

In the adaptive fitness group, we saw much smaller activation increases in the left supramarginal gyrus and superior parietal lobe compared to baseline during the *whole-body imagery fMRI task* (**Fig. 6B**, **Table 3**).

## 4 Discussion

In this pilot clinical trial, our aims were to determine if 8 weeks of CMR or adaptive fitness (1) improved sensory and motor function (primary) and reduced neuropathic pain when present (secondary), and (2) enhanced brain activity and connectivity related to sensorimotor function and body awareness.

The results provide promising preliminary evidence that CMR may address the dual burden of sensorimotor deficits and chronic neuropathic pain in adults with SCI. Both interventions yielded clinical benefits, but CMR produced greater sensorimotor function improvements (reflected by large effect sizes for AIS and NRS) than fitness. These gains should be interpreted in light of injury variability. For instance, upper limb NRS effect sizes were constrained by the fact that 8 participants (7 with paraplegia, 1 with quadriplegia) achieved a maximum score for this item at baseline. The remaining two participants with paraplegia did not reach maximum scores on NRS upper limb function due to a right frozen shoulder in one, and a high thoracal lesion (T4) limiting trunk balance during upper limb activities in the other.

Both groups showed large PSFS effect sizes and improvements on nearly all SCI-FI/AT measures for adults with quadriplegia. For adults with paraplegia, self-reported ease of performing daily functions was markedly better in the CMR group vs the fitness group. The CMR group experienced large improvements for basic mobility, selfcare, and fine motor function, and moderate improvement for ambulation across both time points. The fitness group showed more varied results, with no lasting basic mobility improvements, moderate self-care and fine motor function gains at follow-up, and large ambulation gains at follow-up.

These results show the importance of using both objective and subjective measures when assessing motor function. Objective measures do account for adaptive equipment use, whereas self-reported daily function does, which can produce divergent results. For example, the participants in the fitness group did not improve in objective measures as much as the CMR group for lower limb function; yet, participants with paraplegia self-reported on average that walking with adaptive equipment became easier after the fitness intervention and at follow-up.

Sensory function also improved more after CMR than after adaptive fitness. Given how closely MBR is integrated with sensorimotor function, it follows that both sensory and motor function improve when whole-body awareness, awareness of the relationship between body parts, and spatial body awareness are restored. Across our pilot studies, CMR consistently produces both sensory and motor function gains, which is encouraging, as sensory recovery has been difficult to achieve with other neurorehabilitation treatments.

To date, only one recent study of D’Amico et al. (2026)(63) examined the effects of cervical transcutaneous spinal cord stimulation combined with rehabilitation, finding that 55% of participants with chronic SCI increased total AIS sensory scores by more than 5.19 [the minimum clinically important difference, MCID, for acute and subacute SCI(64) populations; there is no current MCID available for chronic SCI].(63) Their study design did not include a control group or follow-up measures.(63) In comparison, our CMR group improved total AIS sensory scores by more than 7 points, at both post-intervention and at 3-months follow-up: touch improved post-CMR by 7.50±4.88 and 8.06±4.66 points at follow-up. Pinprick improved by 8.50±4.64 and 9.88±4.78 points, respectively. These are the largest improvements we have encountered in literature, sustained for at least 3 months. Notably, direct score comparisons are limited by differences in injury profile: D’Amico et al. (2026)(63) enrolled only participants with cervical incomplete lesion (with majority C4-C6; AIS C-D), whereas our cohort included 44% cervical incomplete lesions (C1-C7; majority of this subgroup AIS D). Nonetheless, our prior study in 26 adults with SCI-related neuropathic pain (where 53% had AIS A) demonstrated that CMR is also effective in persons with complete SCI (AIS A).(16) In our present study, fitness improved touch by 2.14±4.38 and 2.50±4.47 points; and pinprick 2.00±3.81 and 3.50±4.72 points, at the two timepoints.

Furthermore, CMR produced consistent small-to-moderate reductions in neuropathic pain and symptom severity (DN4) while the fitness group showed no change or slight exacerbation of these symptoms (except for a small decrease in average pain). With only 10 participants with pain (5 per group) these findings are preliminary. However, our prior study in 26 adults with SCI-related neuropathic pain demonstrated significant pain reduction after 6 weeks of CMR (highest NPRS: almost 5-point reduction; average NPRS: 4-point reduction; 35% pain-free) sustained at 1-year follow-up in a subgroup of 14 participants who agreed to be tested.(16)

Clinical improvements observed after CMR coincided with enhanced brain connectivity and activation. Our earlier study demonstrated that adults with SCI have weaker connectivity networks and less brain activation in areas responsible for sensorimotor function and MBR than healthy adults.(9) In this study, following 8 weeks of CMR, resting-state connectivity strengthened between the right parietal operculum and right precentral gyrus; and between the right posterior cingulate cortex (as region-of-interest) and the left precentral gyrus, left insula, and left parietal operculum (parts OP1/OP4).(65,66)

The posterior cingulate cortex plays a key role in self-localization, as identified in an out-of-body fMRI study,(11) though the intraparietal-retrosplenial-hippocampal connections appear more relevant in that context. Given our small sample size, we cautiously speculate that the observed connectivity seen in our study between the posterior cingulate cortex and precentral cortex represents self-localization of the ‘actual body’/‘of the self’ being linked to motor areas to guide movements, analogous to how the parietal operculum, insula, and posterior parietal cortex collects, integrates, and transmits multisensory information for motor guidance.

*Imagery of feeling the left leg* increased activation in the pre– and postcentral gyri and superior parietal lobe, while the *whole-body imagery task* activated the right pre– and postcentral gyrus, bilateral supramarginal gyrus, superior parietal lobe, and angular gyrus to a greater extent post-CMR vs baseline. Consistent with our prior CMR brain imaging findings, the brain not only processes and interprets sensory information from the affected areas, but integrates multisensory and visuospatial information into a coherent conscious spatial body awareness to accurately guide movements.(4,8–10,67)

The fitness group showed markedly smaller activation increases compared to baseline, limited to the left supramarginal gyrus and superior parietal lobe during *whole-body imagery*.

While causal relationships cannot be established, the concurrent improvements in MBR-related networks and clinical outcomes support our central hypothesis that targeting MBR can address both sensorimotor deficits and neuropathic pain. SCI-related sensory deficits create a mismatch between expected and received sensations in the brain, disrupting MBR.(3–5) Given the brain is wired for survival, when unable to locate the (paralyzed) body parts, the brain signals this incongruence through spasms and neuropathic pain. We consistently observe that when participants regain awareness of their body parts’ location, spasms resolve and neuropathic pain diminishes.

Surveys confirmed that body awareness and imagery improved more with CMR than fitness. For example, at 3-month follow-up, the CMR group showed moderate-to-large effect sizes for BARQ-R and MAIA-2’s subdomains of noticing, not worrying, emotional awareness, self-regulation and body listening─no moderate or large effect sizes were seen in the fitness group. This confirms prior findings in a larger SCI cohort showing improved BARQ-R (n=69), and MAIA-2’s subscale ‘not worrying’ (n=43) after CMR or Qigong.(68) By helping participants correctly identify sensations of body positions, touch, pressure, and other multisensory variables, CMR reduces the brain’s alarm response: sensations are recognized rather than flagged as unknown, diminishing neuropathic pain signaling.(69)

Participants reported that they were more attuned to their body signals, and reacted to them faster, e.g., when they felt they had a urinary tract infection. Nevertheless, none of the scales related to body awareness or interoception directly assessed SCI-specific MBR. We are currently developing a new SCI-BodyMap to address this gap.(70,71)

The clinical and neuroimaging findings parallel participant reports of increased body awareness after CMR─such as being able to accurate sense where there paralyzed limbs were in space, as well as the contact of the soles of the feet with the floor─which subsequently facilitated transfers and activities of daily living. Crucially, reports from participants in the CMR group encompassed benefits across several domains: sensory, motor, autonomic, and cognitive function. Here are some examples below:

[woman in her 50s] “*I think of parts of my body that I have not thought about in over 20 years and it’s interesting. I find myself touching my feet and legs a bit differently and recognizing they are in fact still there. Sensation in the feet returns, also sensation on the soles of the feet. Transfers feel easier*.”

[man in his 50s] “*The right big toe has woken up, and I have more and more ongoing sensation in the buttocks*. *This is different from other therapies. I am now able to get out of a low sports car without help. My hips move better and feel more free. I am able to get in the RV up the stairs and then sit down with weight equal[ly distributed] in my feet. I continue to walk faster with smart crutches*.”

[man in his 30s] “*My right foot is not dragging anymore. I use a new way of walking now, more natural, faster and smoother, and I am not fatigued. I stand more on top of my feet. It was a no-brainer that it [CMR] was worth it*.”

[man in his 30s] “*I used to lean on the right side all the time, now I am sitting straight. My left hand function is better now. I can hold the bottle with my left hand; I could not do that as well before. My handgrip is normal now. Before it would slip (grip was too weak) and that is not happening now. I stood up with a walker; I could not do that before for more than 20 years. Transfers are better. Before I could only transfer from right to left and now, I can also transfer from left to right. Neuropathy is half of what it used to be.”*

[woman in her 60s] “*For the first time I was able to touch my left foot with my hand in crossed leg position, without back support by leaning my trunk a little forward and using the right foot as a support*.”

[[man in his 30s] “*It is an interesting journey. The CMR helps me also with my depression and clear-thinking problem. I am at a point where I start getting more clarity, feeling awake, and clear thinking, like a runner’s high. I no longer feel “crushing” pain on the right side. This week, I could feel my cat on my body on the left side during the night in an area I am usually not able to feel*.”

[man in his 50s]: *“I did not know that internal map was incongruent with reality; it’s very revealing; a 180-degree paradigm shift. Fine tuning what needs to be finetuned. Very interesting stuff. I had no clonus in the left leg during sessions last week. I do not have restless leg syndrome anymore at night, and that used to wake me up before. I can now transfer from a lower surface to my chair whereas previously, the treatment table needed to be significantly higher. Overall, transfers are better now.”*

[woman in her 50s] *“(week 2): I can cut my nails with both hands now which I was not able to do before. (week 5): all my goals are reached: I can now bike 12 miles, clip toenails (before it would slip), I can climb a ladder, I can snap with both hands (left better than right). Feet and hands are not going numb anymore, and at night when I lay on my side, my legs do not go numb anymore. My hands and feet do not feel cold anymore*.”

Participants also applied what they learned in CMR to daily life, such as thinking about being grounded; knowing where the feet are; improved transfers and body weight shifting; greater attentiveness to leg and foot sensations; better awareness of when to reposition in the wheelchair.

For the fitness group, feedback included noticeable balance improvements, which helped with walking, increased strength, having more energy, and easier transfers. One person would have preferred one-on-one workouts with a personal trainer (instead of oversight by a trainer), but overall, the group enjoyed working out in the adaptive fitness gym.

Our findings must be interpreted within several limitations. Most importantly, the small sample size(1) precludes definitive efficacy claims and warrants a larger controlled trial, particularly for neuropathic pain, given the wide variability in the present cohort; (2) prevented between-group neuroimaging comparisons and mediation analyses to establish causal brain-outcome links; and (3) precluded subanalyses by injury level (paraplegia vs quadriplegia) or other SCI-related variables. Regarding health disparities, 25% were of minority race, none were veterans, 19% lived rurally, 38% were women, and 25% had significant economic distress or income below the official poverty level. While we achieved some satisfactory levels of recruitment of patients with healthy disparities, we aim to recruit more veterans in future studies, given their higher prevalence of SCI compared to community members (in 2019, 42,000 or about 14% of all adults with SCI were veterans, numbers that are likely higher today).(72,73) A remotely delivered CMR version could also extend reach to more participants living in rural locations.

Despite these limitations, the large CMR effect sizes for objectively assessed sensorimotor recovery (AIS and NRS), alongside indications of reduced neuropathic pain and enhanced body awareness are clinically noteworthy. The small-to-moderate effect sizes in the adaptive fitness group support its viability as active control in a larger trial. Notably, this study demonstrated potential for improvements in sensation and movement even up to 29 years after SCI, consistent with our prior work showing gains up to 56 years post-SCI.(16)

Villiger et al. (2013)’s pilot study with virtual reality-augmented lower limb training combining action observation and training, reported reduced neuropathic pain beyond MCID in 5 of 9 adults with incomplete SCI (ASIA D).(74) While mirror therapy and augmented reality with mirror therapy, have been explored to create a perceptual illusion of movement (e.g. walking) for pain relief with varying results,(75–77) to our knowledge, CMR is the only therapeutic approach that *consciously* targets MBR restoration using the participants’ own body parameters, with consistent evidence of increased brain function after CMR. The therapist’s guidance in restoring MBR is *central* to these gains. Together, these preliminary results indicate that targeting maladaptive cortical changes may yield greater dual-burden recovery than standard adaptive fitness, strongly warrant larger randomized controlled efficacy trials.

## 6 Author Contributions

Every author contributed to the study: Conceptualization (AVdW, LM); Funding acquisition (AVdW); Data curation (AVdW, SC, WD); Formal analysis and data interpretation (AVdW, LZ, TH, KL, BM, LM); Cognitive Multisensory Rehabilitation intervention (SB, JB); Recruitment (AVdW); Data collection (AVdW, SC, WD, BM); Supervision (AVdW); Validation (AVdW, SC, WD, LZ); Visualization (AVdW, LZ); Consultant with lived experience of SCI (SMS); PM&R screening for adaptive fitness readiness (RN); SCI study doctor (LM); Writing – original draft (AVdW); Writing – review & editing (AVdW, SC, SB, JB, WD, LZ, TH, BM, SMS, RN, LM, KL). We confirm that more than one author has directly accessed and verified the underlying data reported in the manuscript (AVdW, SC, JB, LZ).

## 8 Acknowledgments

We thank Dr. Marina Zernitz, Director of the Centro Studi di Riabilitazione Neurocognitiva – Villa Miari (Study Center for Cognitive Multisensory Rehabilitation), Italy, for her consultancy with the content of the CMR training protocol. We would like to thank the MRI technicians Wendy Elvendahl and Matthew White for their help with data acquisition. We would like to extend our profound thanks to Marc Noël for the critical review of the manuscript.

## 9 Data Availability Statement

The datasets generated for this study can be found in the Dryad repository, DOI: 10.5061/dryad.v6wwpzhbb upon publication of the manuscript in a peer-reviewed journal.

## List of acronyms

AIS: ASIA Impairment Scale
ANS: Autonomic Standards Assessment form
BARQ-R: Revised Body Awareness Rating Questionnaire
CHART-SF: Craig Handicap Assessment and Reporting Technique-Short Form
CMR: Cognitive Multisensory Rehabilitation
DN4: Douleur Neuropathique 4
FAS: Functionality Appreciation Scale
fMRI: functional Magnetic Resonance Imaging
ISNCSCI: International Standards for Neurological Classification of Spinal Cord Injury
KVIQ: Kinesthetic and Visual Imagery Questionnaire
MAIA-2: Multidimensional Assessment Interoceptive Awareness – version 2
MBR: Mental Body Representations
MCID: Minimum Clinically Important Difference
MSES: Moorong Self-Efficacy Scale
NPRS: Numeric Pain Rating Scale
PAS: Postural Awareness Scale
PHQ-9: Patient Health Questionnaire
PSFS: Patient-Specific Functional Scale
PSQI: Pittsburgh Sleep Quality Index
SAI/TAI: State Trait Anxiety Inventory
SCI: Spinal Cord Injury
SCI-FI/AT: Spinal Cord Injury Functional Index/Assistive Technology
SCI-QOL: Spinal Cord Injury Quality of Life measurement system
TSK: Tampa Scale for Kinesiophobia
VA: Veterans Affairs

## Figure legends

**Supplementary Table 1.** Demographic and clinical characteristics of adults with spinal cord injury.

|  | **Adults with SCI**<br>**(n=16)** |
| --- | --- |
| Age, years, mean±SD, range (years) | 51.63±8.00<br>(38-64) |
| Sex assigned at birth, n (%)<br>Male<br>Female | 10 (62.50)<br>6 (37.50) |
| Gender Identity, n (%)<br>Male<br>Female<br>Other | 10 (62.50)<br>6 (37.50)<br>0 (0.00) |
| Ethnicity, n (%)<br>Hispanic<br>Non-Hispanic | 0 (0.00)<br>16 (100.00) |
| Race, n (%)<br>White<br>Asian<br>African American/Black<br>Multi-racial<br>Hawaiian or Other Pacific Islander<br>Other | 12 (75.00)<br>0 (0.00)<br>4 (25.00)<br>0 (0.00)<br>0 (0.00)<br>0 (0.00) |
| Veteran, n (%)<br>Yes<br>No | 0 (0.00)<br>16 (100.00) |
| Location of residence, n (%)<br>Rural<br>City or suburb | 3 (18.75)<br>13 (81.25) |
| Marital status |  |
| Single | 5 (31.25) |
| Living with significant other or partner | 0 (0.00) |
| Married | 7 (43.75) |
| Separated | 1 (6.25) |
| Widowed | 0 (0.00) |
| Divorced | 3 (18.75) |
| Other | 0 (0.00) |
| Unknown | 0 (0.00) |
| Baseline neuropathic pain intensity level experienced in the prior week, mean±SD, scored between 0-10 (n=10) | |
| High | 6.30±2.41 |
| Average | 3.80±3.05 |
| Low | 0.20±0.42 |
| Mini-Mental State Examination-Short Form (MMSE-SF), mean±SD, scored between 0-16 | 15.19±0.91 |
| Time since spinal cord injury, years, mean±SD, range (years) | 12.44±10.28<br>(1-29) |
| Etiology of SCI, n (%) |  |
| Traumatic |  |
| Motorized accident (motorcycle, car, snowmobile, plane) | 10 (62.50) |
| Bike accident | 1 (6.25) |
| Fall from greater than standing height | 2 (12.50) |
| Gunshot wound | 2 (12.50) |
| Non-traumatic |  |
| Complications from surgery | 1 (6.25) |
| Spinal cord injury lesion level, n (%) |  |
| Cervical (C3-C7) | 7 (43.75) |
| Thoracal (T4-T12) | 6 (37.50) |
| Lumbar (L1-L3) | 3 (18.75) |
| Sacral | 0 (0.00) |
| Type of paralysis |  |
| Quadriplegia | 6 (37.50) |
| Brown-Sequard | 1 (6.25) |
| Paraplegia | 9 (56.25) |
| AIS level, n (%) |  |
| A | 1 (6.25) |
| B | 4 (25.00) |
| C | 4 (25.00) |
| D | 7 (43.75) |
| Ambulation, n (%) |  |
| Walking without assistance | 2 (12.50) |
| Combined use of walking with and without assistance | 1 (6.25) |
| Walking with assistance | 0 (0.00) |
| Combined use of walking with assistance and manual wheelchair | 2 (12.50) |
| Manual Wheelchair | 6 (37.50) |
| Combined use of Manual and Motorized Wheelchair | 2 (12.50) |
| Motorized Wheelchair | 3 (18.75) |
| Neuropathic pain medication taken at baseline, n (%) (n=5) |  |
| Amitriptyline | 1 (20.00) |
| Baclofen | 1 (20.00) |
| Gabapentin | 3 (60.00) |
| Hydrocodone | 1 (20.00) |
| Lyrica | 1 (20.00) |
| Medical cannabis (CBD/THC) | 1 (20.00) |

**Supplementary Table 2.** Primary and secondary outcome measures at three time points.

| Outcome measures | Time point 1 |  | Time point 2 |  | Time point 3 |  | Effect size |  | Effect size |  |
| --- | --- | --- | --- | --- | --- | --- | --- | --- | --- | --- |
|  | Baseline<br>(mean±SD) |  | Post-intervention<br>(mean±SD) |  | Post-CMR<br>(mean±SD) |  | Pre – Post |  | Pre – 3MFU |  |
|  | CMR group | Adaptive<br>Fitness group | CMR group | Adaptive<br>Fitness group | CMR group | Adaptive<br>Fitness group | CMR<br>group | Adap.<br>Fitness<br>group | CMR<br>group | Adap.<br>Fitness<br>group |
| n | (n=8) | (n=7) | (n=8) | (n=7) | (n=8) | (n=6) |  |  |  |  |
| **Highest NPRS** | 3.38±3.29 | 5.14±4.02 | 2.50±2.33 | 5.43±3.64 | 2.00±2.45 | 4.33±3.72 | 0.48 | -0.19 | 0.63 | 0.00 |
| **Average NPRS** | 1.50±2.51 | 3.71±3.40 | 0.38±0.74 | 3.57±3.46 | 0.63±1.19 | 2.83±2.40 | 0.46 | 0.38 | 0.35 | 0.41 |
| **Lowest NPRS** | 0.25±0.71 | 0.29±0.49 | 0.25±0.71 | 0.29±0.49 | 0.25±0.71 | 0.33±0.52 | 0.00 | 0.00 | 0.00 | 0.00 |
| **DN4** | 3.50±2.67 | 4.57±2.64 | 2.88±2.10 | 4.71±2.63 | 2.88±2.10 | 5.00±2.76 | 0.59 | -0.38 | 0.59 | -0.41 |
| n | (n=8) | (n=7) | (n=8) | (n=7) | (n=8) | (n=6) |  |  |  |  |
| **AIS exam** |  |  |  |  |  |  |  |  |  |  |
| - sensory touch (Avg L, R) | 42.69±7.69 | 44.57±5.66 | 50.19±4.99 | 46.71±6.41 | 50.75±4.53 | 45.50±5.74 | 1.54 | 0.49 | 1.73 | 0.56 |
| - sensory pin (Avg L, R) | 40.00±6.44 | 40.79±8.66 | 48.50±6.08 | 42.79±7.89 | 49.88±4.85 | 42.08±5.83 | 1.83 | 0.53 | 2.07 | 0.74 |
| - lower limb motor function | 9.31±8.29 | 12.64±7.99 | 12.69±8.23 | 13.79±8.69 | 13.00±8.55 | 14.58±9.35 | 1.32 | 0.74 | 1.28 | 0.63 |
| **NRS** |  |  |  |  |  |  |  |  |  |  |
| -core | 27.00±11.66 | 35.00±12.78 | 31.38±11.08 | 36.57±13.26 | 33.13±11.58 | 39.17±12.94 | 2.19 | 0.73 | 1.74 | 0.62 |
| -arm function | 135.75±9.84 | 131.00±19.86 | 141.75±5.26 | 131.86±20.12 | 142.63±3.89 | 131.17±19.44 | 0.69 | 0.34 | 0.79 | 0.85 |
| -standing | 15.13±10.56 | 15.43±12.35 | 16.88±11.96 | 15.43±12.35 | 17.13±11.98 | 17.00±13.56 | 0.74 | 0.00 | 0.78 | 0.65 |
| **PSFS** | 2.00±0.84 | 2.24±2.11 | 7.46±2.65 | 4.00±2.36 | 7.79±2.82 | 6.56±1.75 | 1.99 | 1.07 | 1.95 | 1.81 |
| n | (n=5) | (n=4) | (n=5) | (n=4) | (n=5) | (n=3) |  |  |  |  |
| **SCI-FI/AT paraplegia** |  |  |  |  |  |  |  |  |  |  |
| - basic mobility | 29.20±3.70 | 25.75±11.30 | 33.40±2.41 | 26.50±10.97 | 33.60±2.30 | 29.33±11.55 | 1.47 | 0.78 | 1.37 | 0.00 |
| - self-care | 23.20±5.81 | 28.75±6.29 | 30.20±6.65 | 28.50±6.61 | 30.20±6.65 | 31.67±4.51 | 1.41 | -0.50 | 1.41 | 0.58 |
| - fine motor function | 29.00±2.55 | 30.00±4.00 | 30.80±1.79 | 30.00±4.00 | 31.00±1.41 | 30.33±2.89 | 1.38 | 0.00 | 1.27 | 0.58 |
| - ambulation | 5.00±6.86 | 4.50±5.45 | 5.80±7.95 | 5.00±6.27 | 5.80±7.95 | 7.67±7.09 | 0.61 | 0.50 | 0.61 | 1.09 |
| n | (n=3) | (n=3) | (n=3) | (n=3) | (n=3) | (n=3) |  |  |  |  |
| **SCI-FI/AT quadriplegia** |  |  |  |  |  |  |  |  |  |  |
| - basic mobility | 32.67±4.93 | 33.00±3.61 | 33.33±4.62 | 34.33±2.08 | 33.33±4.62 | 34.33±2.08 | 1.16 | 0.87 | 1.16 | 0.87 |
| - self-care | 35.33±0.58 | 33.33±4.62 | 36.00±0.00 | 33.67±4.04 | 36.00±0.00 | 33.67±4.04 | 1.16 | 0.58 | 1.16 | 0.58 |
| - fine motor function | 32.00±0.00 | 28.67±1.15 | 32.00±0.00 | 30.67±2.31 | 32.00±0.00 | 30.67±2.31 | 0.00 | 1.00 | 0.00 | 1.00 |
| - ambulation | 7.67±11.59 | 11.67±10.21 | 8.67±13.32 | 14.00±12.17 | 8.67±13.32 | 14.00±12.17 | 0.58 | 1.12 | 0.58 | 1.12 |
| n | (n=8) | (n=7) | (n=8) | (n=7) | (n=8) | (n=6) |  |  |  |  |
| **Spasms** |  |  |  |  |  |  |  |  |  |  |
| - frequency | 0.25±0.46 | 1.00±0.82 | 0.00±0.00 | 1.00±0.82 | 0.00±0.00 | 0.67±0.82 | 0.54 | 0.54 | 0.00 | 0.41 |
| - severity | 0.50±1.07 | 1.00±0.82 | 0.00±0.00 | 0.86±0.69 | 0.00±0.00 | 0.67±0.82 | 0.47 | 0.38 | 0.47 | 0.65 |
| PSQI | 9.50±4.14 | 6.57±5.00 | 8.00±4.96 | 5.71±5.06 | 9.00±6.07 | 4.67±3.88 | 0.34 | 0.31 | 0.12 | 0.22 |
| Trait Anxiety Inventory | 35.25±15.66 | 37.14±9.67 | 33.25±15.27 | 34.43±9.32 | 32.38±15.00 | 32.83±7.88 | 1.53 | 0.56 | 1.59 | 0.72 |
| State Anxiety Inventory | 26.88±8.46 | 29.86±10.30 | 28.25±8.84 | 29.86±7.67 | 26.25±8.96 | 32.67±7.17 | -0.35 | 0.00 | 0.13 | -0.64 |
| PHQ-9 | 3.88±7.04 | 3.71±2.93 | 4.25±7.50 | 4.14±3.98 | 4.50±7.37 | 3.50±3.27 | -0.29 | -0.25 | -0.68 | 0.10 |
| TSK | 33.50±7.69 | 33.00±5.97 | 35.63±8.05 | 31.71±2.75 | 31.00±8.00 | 28.83±4.79 | -0.77 | 0.32 | 0.75 | 0.78 |
| ANS | 31.38±6.97 | 33.29±6.82 | 32.00±7.01 | 32.86±6.77 | 32.00±7.01 | 32.83±6.18 | 0.68 | -0.55 | 0.68 | -0.18 |
| MSES | 90.38±22.46 | 97.86±13.52 | 88.63±22.28 | 96.57±20.10 | 92.50±19.18 | 95.83±11.79 | -0.27 | -0.17 | 0.25 | -0.17 |
| FAS | 3.02±0.89 | 3.18±0.75 | 3.46±0.63 | 3.14±0.75 | 3.37±0.52 | 3.24±0.61 | 0.61 | -0.13 | 0.52 | 0.10 |
| PAS | 44.13±12.35 | 50.71±6.40 | 51.13±10.09 | 55.57±8.62 | 46.75±11.40 | 52.83±6.18 | 0.60 | 0.54 | 0.25 | 0.34 |
| BARQ-R | 17.63±5.58 | 17.00±6.35 | 17.75±4.83 | 17.71±4.23 | 15.63±6.14 | 16.17±4.75 | -0.02 | -0.17 | 0.59 | 0.43 |
| **MAIA-2** |  |  |  |  |  |  |  |  |  |  |
| - noticing | 3.63±0.99 | 3.50±0.69 | 3.75±0.80 | 4.18±0.40 | 3.84±1.09 | 3.54±0.56 | 0.14 | 0.77 | 0.78 | 0.11 |
| - not distracted | 1.52±1.08 | 1.36±1.05 | 1.63±0.79 | 1.88±1.27 | 1.60±0.90 | 1.42±0.75 | 0.11 | 0.34 | 0.11 | 0.04 |
| - not worrying | 2.50±1.06 | 2.86±0.51 | 2.78±0.38 | 2.63±0.27 | 3.13±0.52 | 2.77±0.50 | 0.29 | -0.54 | 0.61 | -0.24 |
| - attention regulation | 3.88±0.89 | 3.69±1.03 | 3.50±0.83 | 3.20±0.99 | 3.75±0.99 | 3.02±0.64 | -0.42 | -0.48 | -0.12 | -0.66 |
| - emotional awareness | 2.93±1.44 | 2.91±1.60 | 3.20±0.76 | 2.69±1.35 | 3.38±1.43 | 2.73±0.78 | 0.25 | 0.25 | 0.57 | 0.15 |
| - self-regulation | 3.06±1.75 | 3.29±0.91 | 3.28±1.41 | 2.61±0.72 | 3.41±1.61 | 2.88±0.52 | 0.25 | -1.37 | 0.86 | -0.29 |
| - body listening | 1.83±1.11 | 2.62±1.42 | 2.21±1.31 | 1.62±1.42 | 2.58±1.24 | 1.78±0.66 | 0.43 | -1.06 | 1.14 | -0.42 |
| - trusting | 3.21±1.67 | 3.86±1.41 | 3.46±1.56 | 3.38±1.34 | 3.46±1.69 | 3.50±1.38 | 0.39 | -0.38 | 0.35 | -0.33 |
| KVIQ | 44.75±4.10 | 45.14±7.13 | 48.38±2.00 | 45.71±7.41 | 48.88±1.73 | 44.83±7.81 | 1.02 | 0.73 | 1.07 | -0.60 |
| **SCI-QOL (T-scores)** |  |  |  |  |  |  |  |  |  |  |
| - positive affect | 55.51±9.16 | 56.47±7.11 | 51.84±6.59 | 55.89±9.65 | 54.80±8.75 | 52.77±6.26 | -0.58 | -0.14 | -0.14 | -0.79 |
| - resilience | 50.70±10.02 | 52.93±7.86 | 51.18±8.02 | 51.67±10.48 | 53.93±10.21 | 52.20±8.09 | 0.08 | -0.16 | 0.71 | -0.48 |
| - self-esteem | 30.89±12.18 | 28.90±11.27 | 32.34±9.76 | 31.36±7.47 | 28.80±11.87 | 30.02±8.67 | 0.31 | 0.45 | -0.49 | 0.08 |
| - ability to participate | 46.33±4.89 | 48.24±5.40 | 47.04±4.18 | 46.11±5.73 | 49.04±4.51 | 45.67±1.56 | 0.17 | -0.57 | 0.65 | -0.30 |
| - independence | 44.59±2.45 | 45.56±6.79 | 44.95±1.83 | 44.61±5.06 | 43.50±2.54 | 45.07±3.15 | 0.18 | -0.35 | -0.37 | -0.07 |
| - pain behavior | 51.40±8.85 | 52.51±10.58 | 50.51±8.79 | 49.86±11.87 | 45.85±9.36 | 52.02±8.84 | 0.12 | 0.47 | 0.89 | 0.20 |
| **CHART-SF** |  |  |  |  |  |  |  |  |  |  |
| - physical independence | 97.19±3.85 | 98.71±2.56 | 97.19±3.85 | 98.82±2.59 | 97.19±3.85 | 99.83±0.41 | 0.00 | 0.38 | 0.00 | 0.41 |
| - cognitive independence | 100.00±0.00 | 100.00±0.00 | 100.00±0.00 | 100.00±0.00 | 100.00±0.00 | 100.00±0.00 | 0.00 | 0.00 | 0.00 | 0.00 |
| - mobility | 65.25±26.14 | 76.79±32.98 | 75.38±18.94 | 80.00±34.37 | 77.50±17.33 | 92.33±11.83 | 1.18 | 0.37 | 0.76 | 0.55 |
| - occupation | 65.00±39.65 | 63.93±45.18 | 70.94±33.20 | 66.79±42.32 | 86.09±63.75 | 76.25±37.21 | 0.36 | 0.44 | 0.47 | 0.41 |
| - social interaction | 87.88±18.07 | 83.86±22.03 | 93.50±17.98 | 86.00±19.45 | 93.50±17.98 | 83.67±20.21 | 0.60 | 0.38 | 0.60 | -0.41 |
| - economic self-sufficiency | 78.52±39.77 | 78.29±31.32 | 78.52±39.77 | 78.29±31.32 | 78.52±39.77 | 76.40±33.87 | 0.00 | 0.00 | 0.00 | 0.00 |

**Legend:** ANS: Autonomic Standards Assessment Form; BARQ-R: Revised Body Awareness Rating Questionnaire; CHART-SF: Craig Handicap Assessment and Reporting Technique; DN4: Douleur Neuropathique (Neuropathic pain) 4; FAS: Functionality Appreciation Scale; ISNCSCI-AIS exam: International Standards for Neurological Classification of SCI American Spinal Cord Injury Association Impairment Scale; KVIQ: Kinesthetic and Visual Imagery Questionnaire; MAIA-2: Multidimensional Assessment of Interoceptive Awareness Version 2; MSES: Moorong Self-Efficacy Scale; NPRS: Numeric Pain Rating Scale (used to measure neuropathic pain in the prior week); NRS: Neuromuscular Recovery Scale; PAS: Postural Awareness Scale; PSFS: Patient Specific Functional Scale; SCI: spinal cord injury; PHQ-9: Patient Health Questionnaire-9; PSQI: Pittsburgh Sleep Quality Index; SCI-FI/AT: Spinal Cord Injury Functional Index/Assistive Technology Short Forms; SCI-QOL: Spinal Cord Injury-Quality of Life scales; TSK: Tampa Scale for Kinesiophobia. **Note:** Positive and negative effect size values indicate whether there was improvement or worsening, respectively, of outcome measures. One person in the adaptive fitness group withdrew before the follow-up testing, so averages and SD are calculated n-1 for this time point. Effect sizes pre-3MFU also reflect the reduced n. All *p*-values are adjusted using Benjamini-Hochberg False Discovery Rate (FDR) for multiple comparison corrections

**Supplementary Table 3.**
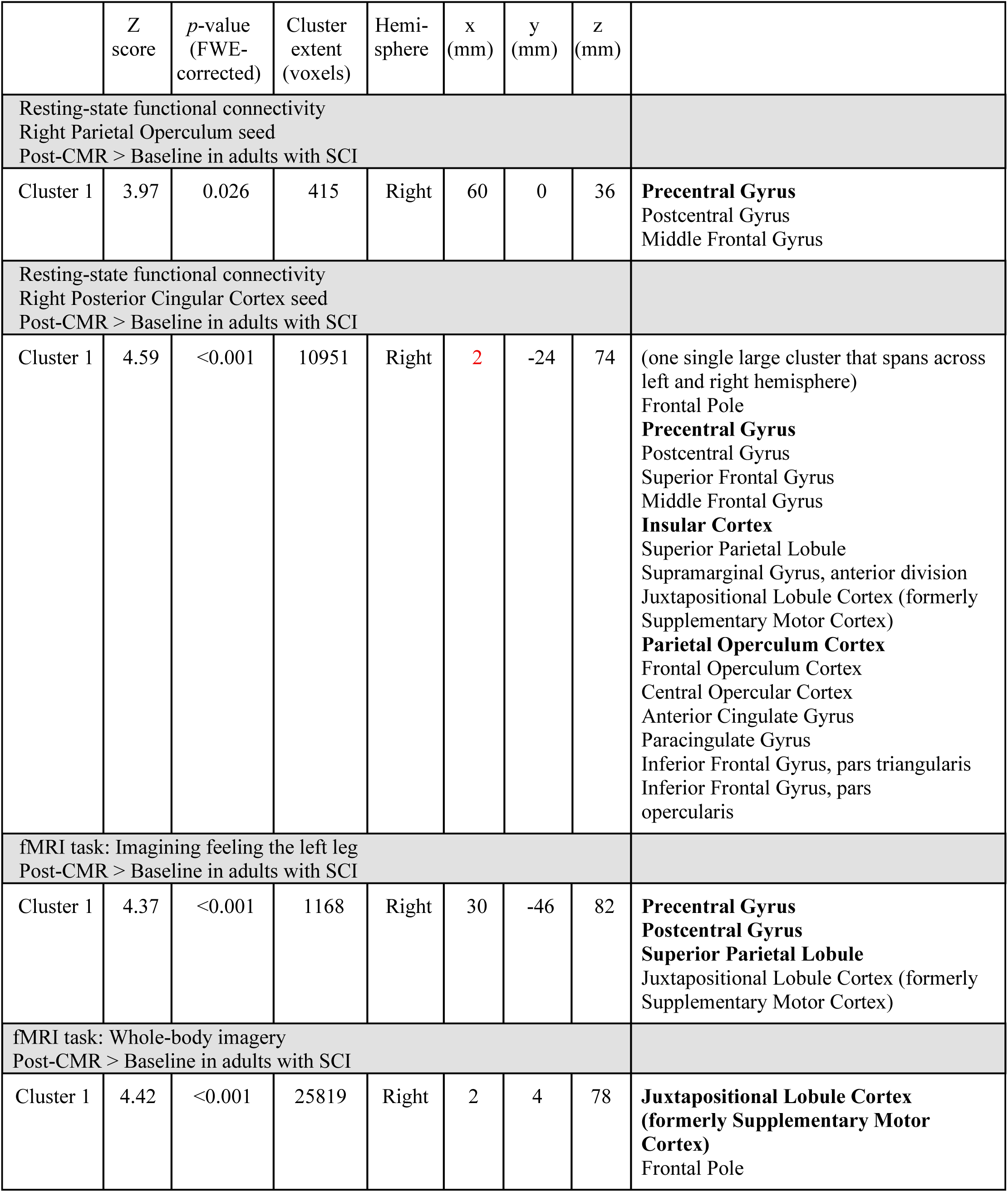
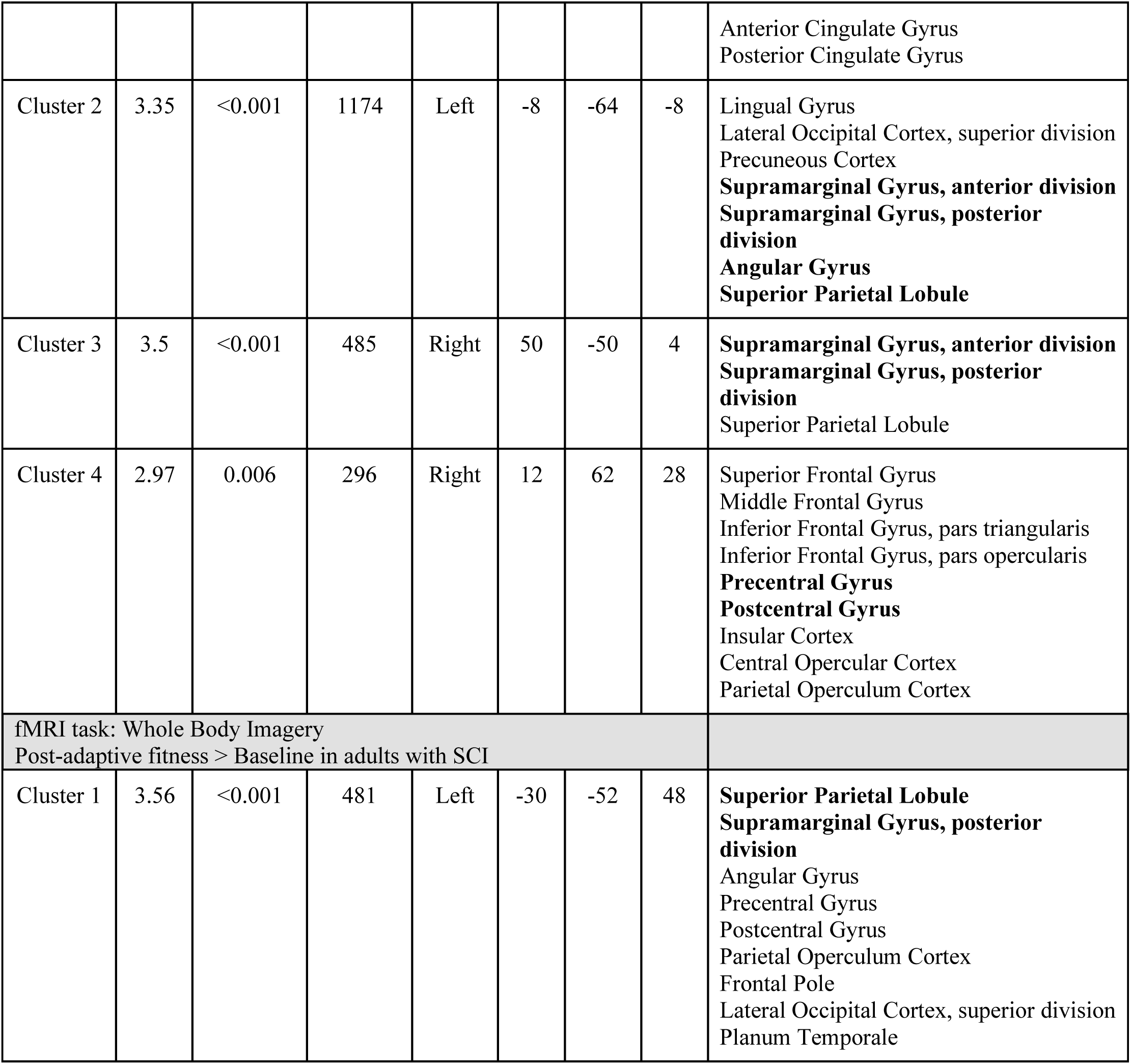
Brain imaging outcomes for resting-state and task-based fMRI in adults with SCI.

|  | Z score | p-value (FWE-corrected) | Cluster extent (voxels) | Hemi-sphere | x (mm) | y (mm) | z (mm) |  |
| --- | --- | --- | --- | --- | --- | --- | --- | --- |
| Resting-state functional connectivity<br>Right Parietal Operculum seed<br>Post-CMR > Baseline in adults with SCI |  |  |  |  |  |  |  |  |
| Cluster 1 | 3.97 | 0.026 | 415 | Right | 60 | 0 | 36 | **Precentral Gyrus**<br>Postcentral Gyrus<br>Middle Frontal Gyrus |
| Resting-state functional connectivity<br>Right Posterior Cingular Cortex seed<br>Post-CMR > Baseline in adults with SCI |  |  |  |  |  |  |  |  |
| Cluster 1 | 4.59 | <0.001 | 10951 | Right | 2 | -24 | 74 | (one single large cluster that spans across left and right hemisphere)<br>Frontal Pole<br>**Precentral Gyrus**<br>Postcentral Gyrus<br>Superior Frontal Gyrus<br>Middle Frontal Gyrus<br>**Insular Cortex**<br>Superior Parietal Lobule<br>Supramarginal Gyrus, anterior division<br>Juxtapositional Lobule Cortex (formerly Supplementary Motor Cortex)<br>**Parietal Operculum Cortex**<br>Frontal Operculum Cortex<br>Central Opercular Cortex<br>Anterior Cingulate Gyrus<br>Paracingulate Gyrus<br>Inferior Frontal Gyrus, pars triangularis<br>Inferior Frontal Gyrus, pars opercularis |
| fMRI task: Imagining feeling the left leg<br>Post-CMR > Baseline in adults with SCI |  |  |  |  |  |  |  |  |
| Cluster 1 | 4.37 | <0.001 | 1168 | Right | 30 | -46 | 82 | **Precentral Gyrus**<br>**Postcentral Gyrus**<br>**Superior Parietal Lobule**<br>Juxtapositional Lobule Cortex (formerly Supplementary Motor Cortex) |
| fMRI task: Whole-body imagery<br>Post-CMR > Baseline in adults with SCI |  |  |  |  |  |  |  |  |
| Cluster 1 | 4.42 | <0.001 | 25819 | Right | 2 | 4 | 78 | **Juxtapositional Lobule Cortex (formerly Supplementary Motor Cortex)**<br>Frontal Pole |
|  |  |  |  |  |  |  |  | Anterior Cingulate Gyrus<br>Posterior Cingulate Gyrus |
| Cluster 2 | 3.35 | <0.001 | 1174 | Left | -8 | -64 | -8 | Lingual Gyrus<br>Lateral Occipital Cortex, superior division<br>Precuneous Cortex<br>**Supramarginal Gyrus, anterior division**<br>**Supramarginal Gyrus, posterior division**<br>**Angular Gyrus**<br>**Superior Parietal Lobule** |
| Cluster 3 | 3.5 | <0.001 | 485 | Right | 50 | -50 | 4 | **Supramarginal Gyrus, anterior division**<br>**Supramarginal Gyrus, posterior division**<br>Superior Parietal Lobule |
| Cluster 4 | 2.97 | 0.006 | 296 | Right | 12 | 62 | 28 | Superior Frontal Gyrus<br>Middle Frontal Gyrus<br>Inferior Frontal Gyrus, pars triangularis<br>Inferior Frontal Gyrus, pars opercularis<br>**Precentral Gyrus**<br>**Postcentral Gyrus**<br>Insular Cortex<br>Central Opercular Cortex<br>Parietal Operculum Cortex |
| fMRI task: Whole Body Imagery<br>Post-adaptive fitness > Baseline in adults with SCI |  |  |  |  |  |  |  |  |
| Cluster 1 | 3.56 | <0.001 | 481 | Left | -30 | -52 | 48 | **Superior Parietal Lobule**<br>**Supramarginal Gyrus, posterior division**<br>Angular Gyrus<br>Precentral Gyrus<br>Postcentral Gyrus<br>Parietal Operculum Cortex<br>Frontal Pole<br>Lateral Occipital Cortex, superior division<br>Planum Temporale |

